# Delineating clinical and developmental outcomes in *STXBP1*-related disorders

**DOI:** 10.1101/2023.05.10.23289776

**Authors:** Julie Xian, Kim Marie Thalwitzer, Jillian McKee, Katie Rose Sullivan, Elise Brimble, Eryn Fitch, Jonathan Toib, Michael C. Kaufman, Danielle deCampo, Kristin Cunningham, Samuel R. Pierce, James Goss, Charlene Son Rigby, Steffen Syrbe, Michael Boland, Ben Prosser, Nasha Fitter, Sarah M. Ruggiero, Ingo Helbig

**Author notes:** Corresponding author: Ingo Helbig, MD Division of Neurology Children’s Hospital of Philadelphia Philadelphia, PA 19104 /.

## Abstract

*STXBP1*-related disorders are among the most common genetic epilepsies and neurodevelopmental disorders. However, the longitudinal epilepsy course and developmental endpoints have not yet been described in detail, which is a critical prerequisite for clinical trial readiness. Here, we assessed 1,281 cumulative patient-years of seizure and developmental histories in 162 individuals with *STXBP1*-related disorders and established a natural history framework. *STXBP1*-related disorders are characterized by a dynamic pattern of seizures in the first year of life and high variability in neurodevelopmental trajectories in early childhood. Epilepsy onset differed across seizure types, with 90% cumulative onset for infantile spasms by 6 months and focal-onset seizures by 27 months of life. Epilepsy histories diverged between variant subgroups in the first 2 years of life, when individuals with protein-truncating variants and deletions in *STXBP1* (n=39) were more likely to have infantile spasms between 5 and 6 months followed by seizure remission, while individuals with missense variants (n=30) had an increased risk for focal seizures and ongoing seizures after the first year. Developmental outcomes were mapped using milestone acquisition data in addition to standardized assessments including the Gross Motor Function Measure-66 Item Set and the Grasping and Visual-Motor Integration subsets of the Peabody Developmental Motor Scales. Quantification of endpoints revealed high variability during the first five years of life, with emerging stratification between clinical subgroups, most prominently between individuals with and without infantile spasms. We found that individuals with neonatal seizures or early infantile seizures followed by seizure offset by 12 months of life had more predictable seizure trajectories in early to late childhood than compared to individuals with more severe seizure presentations, including individuals with refractory epilepsy throughout the first year. Characterization of anti-seizure medication response revealed age-dependent response over time, with phenobarbital, levetiracetam, topiramate, and adrenocorticotropic hormone effective in reducing seizures in the first year of life, while clobazam and the ketogenic diet were effective in long-term seizure management. Virtual clinical trials using seizure frequency as the primary outcome resulted in wide range of trial success probabilities across the age span, with the highest probability in early childhood between 1 year and 3.5 years. In summary, we delineated epilepsy and developmental trajectories in *STXBP1*-related disorders using standardized measures, providing a foundation to interpret future therapeutic strategies and inform rational trial design.

## Introduction

*STXBP1*-related disorders are among the most common genetic epilepsies and neurodevelopmental disorders, with an estimated frequency of 1:30,000.^1, 2^ The phenotypic landscape is characterized by neurodevelopmental delay in 95% and epilepsy in more than 80% of individuals.^3^ However, the wide range of neurodevelopmental features has made it difficult to delineate phenotypic subgroups,^4–9^ map genotype-phenotype associations, and understand treatment response. Accordingly, the significant heterogeneity within *STXBP1*-related disorders has limited understanding of developmental progression and the range of clinical outcomes.

*STXBP1* encodes the syntaxin-binding protein (MUNC18-1), a key organizer of the neuronal SNARE complex, the molecular machinery driving synaptic vesicle fusion.^10, 11^ While the main disease mechanism is haploinsufficiency, dominant negative effects have been suggested as an alternative disease mechanism.^12–14^ Phenotypic signatures associated with specific genetic variants in *STXBP1* have only recently been described,^3^ and efforts to elucidate the genomic architecture and explain the clinical heterogeneity are ongoing. Nevertheless, *STXBP1* represents a focused target for gene therapy and clinical trials, with treatment strategies including chemical chaperones, antisense oligonucleotides, and viral gene therapies on the horizon.^14, 15^

Understanding longitudinal clinical presentations and trajectories in *STXBP1*-related disorders remains an ongoing challenge. First, despite numerous cross-sectional studies on outcomes,^7, 16, 17^ the variability resulting from cohort samples and study design differences has challenged interpretation of findings within and across studies. In addition, larger cohort studies have been limited to broad phenotypic descriptions that did not include longitudinal trajectories.^3^ Furthermore, a major challenge in moving *STXBP1*-related disorders towards clinical trial readiness has been the use of varying outcome measures for clinical endpoints.^18–21^ Lastly, the heterogeneous pattern of seizures limits precise understanding of optimal time windows for clinical trial design.^22, 23^ Accordingly, there remains a critical and unmet need to systematically assess longitudinal clinical data and validate disease and subgroup-specific measures.

To address this gap, we aimed to assess the trajectory of epilepsy and neurodevelopment through validated outcome scales, stratifying trajectories by clinical and genetic subgroups across the age span in individuals with *STXBP1*-related disorders.

## Methods

### Inclusion of individuals with *STXBP1*-related disorders

We assessed the genetic and clinical information of 162 individuals with *STXBP1*-related disorders, including 104 individuals at Children’s Hospital of Philadelphia and 58 additional individuals from the Ciitizen Natural History Study Patient Registry for whom only milestone acquisition data was analyzed in this study. Only variants considered pathogenic or likely pathogenic according to American College of Medical Genetics and Genomics (ACMG) criteria or *de novo* variants were included.

### Reconstruction and assessment of epilepsy and developmental histories

We reconstructed 765 patient-years of epilepsy histories across 101 individuals with *STXBP1*-related disorders for whom detailed seizure information was available in the medical records. Using a previously published outcome scale derived from the Pediatric Epilepsy Learning Health System (PELHS)-championed framework for seizure severity,^3, 24^ we captured >20 seizure types and monthly seizure frequencies (SF), quantified as: multiple daily seizures (>5 per day, SF score = 5), several daily seizures (2–5 per day, SF score = 4), daily seizures (SF score = 3), weekly seizures (SF score = 2), monthly seizures (SF score = 1), and no seizures (SF score = 0). For individuals in which seizures were noted to be present but seizure frequency was unknown, we calculated the median seizure frequency within the cohort during the respective month. Seizure types were captured using the Human Phenotype Ontology (HPO), a controlled dictionary for harmonization of phenotypic data.^25, 26^

To assess development, we included analysis of (a) milestone acquisition and (b) outcomes captured from routine clinical care through validated standardized measures and classification scales. The Gross Motor Function Measure 66 – Item Sets (GMFM-66-IS) was completed by a physical therapist to measure motor function in areas such as rolling, sitting, crawling, standing, walking, running, and jumping.^27^ The Grasping and Visual-Motor Integration subtests of the Peabody Developmental Motor Scales – Second Edition (PDMS-2) was completed by an occupational therapist to assess fine motor function.^28^ Raw scores, percentiles, and age equivalents were calculated for the PDMS-2 while the GMFM-66-Is provided a single interval level score.

Gross motor, fine motor, and language abilities were categorized using five-level classifications systems. The Gross Motor Function Classification System Extended & Revised (GMFCS-ER) describes how children are able to self-initiate movements in activities such as sitting, transfers, and mobility.^21, 29^ The Manual Ability Classification System (MACS) was developed for children aged 4 to 18 years old and describes their ability to use their hands with objects during activities of daily living, with a focus on the use of both hands together.^30^ The Communication Function Classification System (CFCS) describes everyday communication performance including sender roles, receiver role, pace of communication, degree of familiarity with a communication partner, age-appropriateness, and the use of augmentative and alternative communication.^31^ For all classification scales, Level I indicates high levels of function while Level V indicates decreased levels of function. The reliability and content validity of the GMFCS-ER, MACS, and CFCS have been supported in multiple investigations.^30–32^

### Longitudinal seizure frequency forecasting and quantification of seizure predictability

Given the heterogeneity of epilepsy in *STXBP1*, we then aimed to stratify subgroups based on the variability in seizures across the age span. We developed a longitudinal seizure frequency forecasting model to characterize the predictability of the epilepsy course. In brief, we compared seizure frequencies across monthly time intervals between all combinations of individual pairs during the first 12 months of life and derived a measure of phenotypic resemblance in epilepsy histories (**Supplementary Material**). We then predicted each individuals’ epilepsy trajectory after the first year of life based on the known epilepsy histories in the subgroup of 10 individuals that most closely resembled the individual in the first year of life. Forecasted predictions were compared to a distribution of randomly generated seizure frequencies and permutation testing of 100,000 for each individual estimation was performed to evaluate “better than chance forecasting.”

Comparing the difference between predicted and actual seizure trajectories after the first year of life enabled us to identify subgroups defined by seizure predictability (i.e., degree of difference between forecasted and actual seizure frequencies across months). The stratification of subgroups then allowed us to analyze features of seizure histories in the first year of life that were enriched and more commonly observed in individuals with unpredictable seizures later in life compared to individuals with predictable seizure trajectories.

### Comparative effectiveness and characterization of treatment response

Seizure endpoints were analyzed stratified by anti-seizure medications (ASM) and treatment strategies. The rationale behind this analysis was that the effect of a novel drug should have additional benefit over existing treatment strategies. As previously described,^3, 33^ efficacy was characterized by reduction in seizure frequency for short-term effect and maintaining seizure freedom, defined by no seizures across consecutive months, for long-term treatment effect. The relative effectiveness was determined through assessing the months in which periods of seizure reduction or seizure freedom coincided with certain ASMs compared to months in which no change in seizure frequency or a worsening of seizure frequency was noted. We compared treatments across seizure types and characterized the percent reduction in seizures in temporal relation to treatment initiation.

### A framework for virtual clinical trials

Finally, we aimed to identify time windows during which a treatment effect that decreases seizure burden would have the highest probability of being detected. Accordingly, we developed a framework for modeling clinical trials, analyzing different age ranges across 765 total patient-years of epilepsy histories. For each virtual trial, we sampled 20 individuals with ongoing seizures at the trial start and simulated a 6-month and 12-month period of 10%, 15%, and 20% seizure reduction (**Supplementary Material**). The percent reduction in seizures was calculated based on the cumulative sum of seizure frequencies at the start of the window, and the simulated treatment effect across each trial window was performed by decreasing seizure frequencies across the trial period. We used the synthetic control method to statistically evaluate the simulated effect,^34, 35^ comparing the distribution of seizure frequencies following reduction of seizures against the observed distribution of frequencies in the group. We ran 1,000 virtual trials for each month across the age span and defined a novel measure, the Observed Frequency of Trial Success (OFTS), or the proportion of trials out of 1,000 in which a significant effect was detected. This allowed us to analyze optimal windows during which a treatment response would most likely be observed in a real-world trial when using seizure frequency as the primary outcome.

### Statistical analysis and data availability

All computational analyses were performed using the R Statistical Framework.^36^ The Kaplan-Meier estimate was used to measure the probability of seizure risk and seizure remission, defined by at least 12 consecutive months of seizure freedom,^37^ while the log-rank sum test was used to determine significance between subgroups. The Wilcoxon rank sum test was used for statistical comparison of distributions in seizure frequencies between clinical subgroups in addition to distributions of raw scores for the GMFM-66-IS and PDMS-2 in assessing developmental outcomes. Fisher’s exact test was used for assessing phenotypic correlations between clinical and genetic subgroups and comparative effectiveness of treatment strategies. Associations are presented as odds ratios with 95% confidence intervals, correcting for multiple comparisons using a False Discovery Rate (FDR) of 5%. Primary data is available in the Supplementary Material. All code is available at github.com/helbig-lab/STXBP1-NHS.

## Results

### *STXBP1*-related disorders are characterized by neurodevelopmental differences and a dynamic seizure pattern over time

We assessed the clinical landscape across 162 individuals with *STXBP1*-related disorders and reconstructed longitudinal disease histories across a total observation time of 1,281 patient-years. The median observation time was 5.1 years (IQR 2.75 – 11.3 years), with a minimum of 3 months and maximum of 44.2 years (**Table 1**). We reconstructed epilepsy histories across 9,174 total patient months in 101 individuals for whom detailed seizure data was documented. Of these individuals, 72 individuals (71.3%) had a past or current history of seizures while 29 (28.7%) did not have any seizures. More than 98% of individuals in our cohort had developmental delay or intellectual disability, with the first reported indication of delay at a median of 7 months (n=30, IQR 6 – 9 months). The median age of genetic diagnosis was 1.9 years (IQR 6 months – 5.4 years), with the earliest diagnosis at 3 weeks.

**Table 1.** Cohort information on 162 individuals with *STXBP1*-related disorders

| **Demographics** |  |
| --- | --- |
| Male | 81/162 (50.0%) |
| Female | 81/162 (50.0%) |
| **Age distribution** |  |
| Age at assessment (*n*=162) | Median 5.1 years (IQR 2.75 – 11.3 years)<br>Range 3 months – 44.2 years |
| Seizure onset (*n*=72/101) | Median 2 months (IQR 1 – 7 months)<br>Range neonatal onset – 11.1 years |
| Age at genetic diagnosis (*n*=97, data available) | Median 1.9 years (IQR 6 months – 5.4 years)<br>Range 3 weeks – 20.0 years |
| **Genetic and phenotypic information** |  |
| Genetic variants in *STXBP1* |  |
| Missense variants | 46 individuals |
| p.Arg406Cys/His/Ser | 9 individuals |
| p.Arg292Cys/His/Pro | 8 individuals |
| p.Arg551Cys/His | 4 individuals |
| PTV/del <sup>a</sup> | 55 individuals |
| In-frame insertions/deletions | 3 individuals |
| **Epilepsy data** |  |
| Seizure types (*n*=72/101) |  |
| Focal-onset seizures | 48 individuals |
| Neonatal seizures | 31 individuals |
| Infantile/epileptic spasms | 29 individuals |
| Early infantile seizures | 27 individuals |
| **Developmental data** |  |
| Milestones assessment (age range of cohort assessed) |  |
| Head control | 119 individuals (IQR 2.34 – 9.88 years) |
| Roll over | 135 individuals (IQR 2.67 – 10.01 years) |
| Sit unassisted | 133 individuals (IQR 2.13 – 9.29 years) |
| Grasp and hand control | 109 individuals (IQR 2.65 – 10.04 years) |
| Walk assisted or unassisted | 151 individuals (IQR 2.95 – 10.66 years) |
| Few words/sentences | 103 individuals (IQR 2.44 – 11.02 years) |
| Clinical exams <sup>b</sup> (age at assessment) |  |
| GMFM-66-IS | 45 individuals (IQR 1.57 – 5.35 years) |
| GMFCS-ER | 41 individuals (IQR 1.83 – 7.40 years) |
| PDMS-2 | 21 individuals (IQR 1.17 – 3.75 years) |
| MACS | 9 individuals (IQR 2.75 – 9.5 years) |
| CFCS | 23 individuals (IQR 3.0 – 11.75 years) |

<sup>a</sup> Protein-truncating variants (PTV)/del included splice sites, frameshifts, and whole and partial gene deletions. One individual with a synonymous variant predicted to affect splicing was included in PTV/del.
<sup>b</sup> Abbreviations: Gross Motor Function Measure (GMFM-66-IS); Gross Motor Function Classification System Extended & Revised (GMFCS-ER); Peabody Developmental Motor Scales (PDMS-2); Manual Ability Classification System; (MACS), Communication Function Classification System (CFCS).

Seizures were most prominent in the first year of life (**Figure 1A**). The median age of epilepsy onset was 2 months (IQR 1 – 7 months). 61/72 individuals (84.7%) had seizure onset in the first year of life (**Figure 1B**): 31 individuals had neonatal onset, 30 had infantile onset after the first month with 80% cumulative onset by 10 months of life and 90% by 1.6 years. Onset and trajectory of seizures were highly variable, with relatively few months in which the proportion of all individuals with seizures exceeded 50% (**Figure 1A**, inset). The most common seizure types in the overall cohort were focal-onset seizures (n=48, f=0.67) and infantile spasms (n=29, f=0.40), of which 39 individuals and 29 individuals had onset in the first year of life, respectively. Of individuals who had neonatal seizures, 22/31 (71.0%) had focal-onset seizures, 10 (32.3%) developed tonic seizures, and 14 (45.2%) developed spasms following the neonatal period.

**Figure 1.**
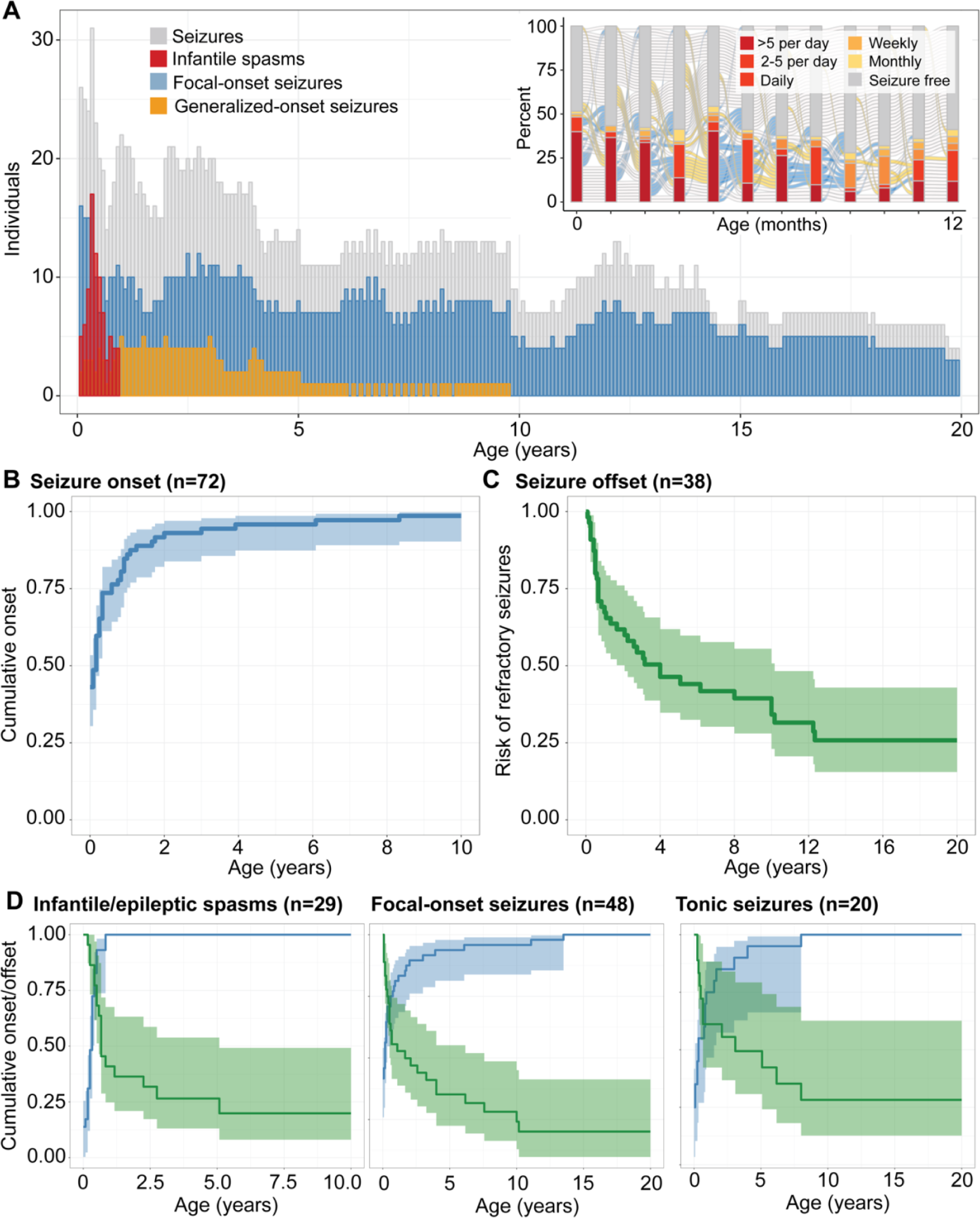
Overall landscape of seizures in *STXBP1*-related disorders. (A) Longitudinal distribution of seizure types across the age span. The heterogeneous pattern of epilepsy progression in the first year of life (inset), showing the proportion of individuals with seizures at a certain frequency during each respective month. **(B)** Cumulative onset of seizures in individuals with epilepsy. **(C)** Seizure remission indicated by risk of having refractory seizures over time, defined by not achieving 12 consecutive months of seizure freedom. **(D)** Stratification by seizure types in *STXBP1* demonstrate a dynamic pattern of risk across the first 10 years of life with infantile spasms most prominent in the first 6 months of life and other seizure types including focal-onset seizures and tonic seizures more broadly distributed.

While seizures were the most prominent in the first year of life, stratification by seizure types demonstrated a dynamic pattern of epilepsy over time (**Figure 1D**; **Table 2**). The risk for infantile spasms was greatest between 2 and 6 months of life (n=29), with a median onset of 5 months and 90% cumulative onset by 9 months. While the seizure onset risk for focal seizures (n=48, median 2 months, IQR neonatal – 7.5 months, 90% cumulative onset by 2.3 years) and tonic seizures (n=20, median 4 months, IQR 0.75 months – 1.53 years, 90% cumulative onset by 3.1 years) were primarily in the first three years of life, the risk for bilateral tonic-clonic seizures presented later in life (n=15, median 3.92 years, IQR 7.5 months – 9.5 years, 90% cumulative onset by 12.2 years). In addition to the age-dependent distribution of seizure onset, 41 (56.9%) individuals presented with more than one seizure type, with infantile spasms (n=22) and focal impaired awareness seizures (n=13) as the most frequent types occurring with at least one additional seizure type.

**Table 2.** Epilepsy and developmental endpoints

| Seizure outcomes |  |  |  |  |
| --- | --- | --- | --- | --- |
| Percentile | 25<sup>th</sup> percentile | 50<sup>th</sup> percentile | 75<sup>th</sup> percentile | 90<sup>th</sup> percentile |
| Seizure onset (n=72) | neonatal onset | 2 months | 7 months | 1.6 years |
| Focal-onset seizures (n=48) | neonatal onset | 2 months | 7.5 months | 2.3 years |
| Infantile/epileptic spasms (n=29) | 1 month | 4 months | 5 months | 6 months |
| Tonic seizures (n=20) | 0.8 months | 4 months | 1.5 years | 3.1 years |
| Bilateral tonic-clonic seizures (n=15) | 7.5 months | 3.9 years | 9.5 years | 12.2 years |
| PTV/del <sup>a</sup> (n=39) | neonatal onset | 2 months | 3 months | 1.7 years |
| Missense variants (n=30) | neonatal onset | 1 month | 6.3 months | 1.1 years |
| Overall seizure remission <sup>b</sup> (n=38/61) | 6 months | 1.2 years | 4.0 years | 10.1 years |
| Focal-onset seizures (n=27) | 3 months | 7 months | 2.9 years | 6.7 years |
| Infantile/epileptic spasms (n=18) | 5 months | 8 months | 1.1 years | 3.5 years |
| Tonic seizures (n=12) | 4.8 months | 8 months | 3.6 years | 6.1 years |
| Bilateral tonic-clonic seizures (n=8) | 1.3 years | 3.6 years | 7.3 years | 8.6 years |
| PTV/del (n=26) | 6 months | 11 months | 4.0 years | 9.0 years |
| Missense variants (n=11) | 7 months | 2.6 years | 5.1 years | 10.2 years |
| Developmental outcomes |  |  |  |  |
| Milestone acquisition |  |  |  |  |
| Head control (n=31) | 3 months | 6.3 months | 1.0 year | 3.0 years |
| Roll over (n=103) | 5 months | 7.6 months | 11 months | 1.2 years |
| Sit unassisted (n=84) | 10 months | 1.0 year | 1.4 years | 2.0 years |
| Grasp and hand control (n=60) | 7 months | 10 months | 1.0 year | 2.0 years |
| Walk assisted or unassisted (n=81) | 1.5 years | 2.4 years | 3.5 years | 5.3 years |
| Few words/sentences (n=22) | 1.6 years | 2.0 years | 3.3 years | 7.0 years |
| Standardized assessments <sup>c</sup> |  |  |  |  |
| GMFM-66-IS (n=45 individuals, 57 exams) |  |  |  |  |
| 0-1 year (n=9 exams) | 19.1 | 25.8 | 27.3 | 29.2 |
| 1 year-2 years (n=15 exams) | 23.7 | 33.9 | 40.2 | 41.9 |
| 2-5 years (n=20 exams) | 25.8 | 35.8 | 46.1 | 61.6 |
| 5+ years (n=13 exams) | 54.9 | 58.9 | 61.2 | 66.6 |
| PDMS-2 (n=21 individuals) |  |  |  |  |
| Grasping (n=23 exams) | 24.5 | 33.5 | 39.0 | 41.8 |
| Visual motor integration (n=23 exams) | 26.5 | 34.5 | 53.0 | 64.5 |
| Classification scales |  |  |  |  |
| GMFCS-ER (n=41) | Level I: n=5, Level II: n=16, Level III: n=4, Level IV: n=12, Level V: n=5 |  |  |  |
| MACS (n=9) | Level I: n=0, Level II: n=4, Level III: n=2, Level IV: n=2, Level V: n=1 |  |  |  |
| CFCS (n=23) | Level I: n=0, Level II: n=1, Level III: n=3, Level IV: n=5, Level V: n=14 |  |  |  |

<sup>a</sup> PTV/del included splice sites, frameshifts, and whole and partial gene deletions.
<sup>b</sup> Seizure remission was defined by at least 12 months of consecutive seizure freedom.
<sup>c</sup> Abbreviations: Gross Motor Function Measure (GMFM-66-IS); Gross Motor Function Classification System Extended & Revised (GMFCS-ER); Peabody Developmental Motor Scales (PDMS-2); Manual Ability Classification System; (MACS), Communication Function Classification System (CFCS).

We then assessed seizure remission, defined as at least 12 consecutive months of seizure freedom. In individuals with at least 1 year of observation time following onset (n=61), 38 individuals had seizure remission, with the median offset at 1.21 years (IQR 6 months – 4.0 years, **Figure 1C**; **Table 2**). The age at offset varied across seizure types, with median offset for focal-onset seizures (n=27) and epileptic spasms at 7 months and 8 months respectively (IQR 3 months – 2.92 years; IQR 5 months – 1.08 years), while the median offset for bilateral tonic-clonic seizures was 3.63 years (IQR 1.31 years – 7.25 years). While seizures reoccurred in 20 individuals following remission, our cohort represented a primarily pediatric population of *STXBP1*-related disorders, and we were not able to make robust conclusions regarding seizure reoccurrence, including childhood to adulthood.

### Epilepsy trajectories diverge when stratifying by genetic variants in *STXBP1*

Given recognition of emerging phenotypic signatures associated with specific variants in *STXBP1*, we then assessed the variant spectrum in our cohort. 46 (44.2%) individuals had missense variants, 15 (14.6%) had frameshift variants, 14 (13.6%) had nonsense variants, 14 had splice site variants (13.6%), 12 (11.7%) had whole or partial gene deletions, and 3 (2.9%) had in-frame insertions or deletions. Individuals with nonsense, splice site, and frameshift variants and whole or partial gene deletions were grouped as PTV/del for further analyses (n=55, 52.9%). Thirty-six individuals had recurrent variants, including the three most frequent genetic hotspots identified in *STXBP1*: p.Arg406Cys/His/Ser (n=9), p.Arg292Cys/His/Leu/Pro (n=8), and p.Arg551Cys/His (n=4). One individual had a novel p.Arg406Ser variant, with no history of epilepsy and a milder clinical presentation compared to the more common p.Arg406Cys/His variants.

First, we delineated the longitudinal epilepsy history in individuals with PTV/del (n=39 with seizures) versus missense variants (n=30 with seizures) (**Figure 2A**). We found the greatest difference in seizure frequency distributions between 10 months and 2 years of life (**Figure 2B**), during which time individuals with missense variants were more likely to have more frequent seizures than individuals with PTV/del. This observed difference was primarily driven by the higher frequency of focal-onset seizures, and a similar effect was also observed between 3.5 to 4 years and 5 to 6 years of life with higher frequencies in focal-onset seizures and tonic seizures associated with missense variants (**Figure 2D**). For individuals with PTV/del, a significant difference was observed between 5 and 6 months of life, when infantile spasms were the most prominent (p=0.03, **Figure 2C**). The risk of intractable seizures was higher in individuals with missense variants (p=0.006).

**Figure 2.**
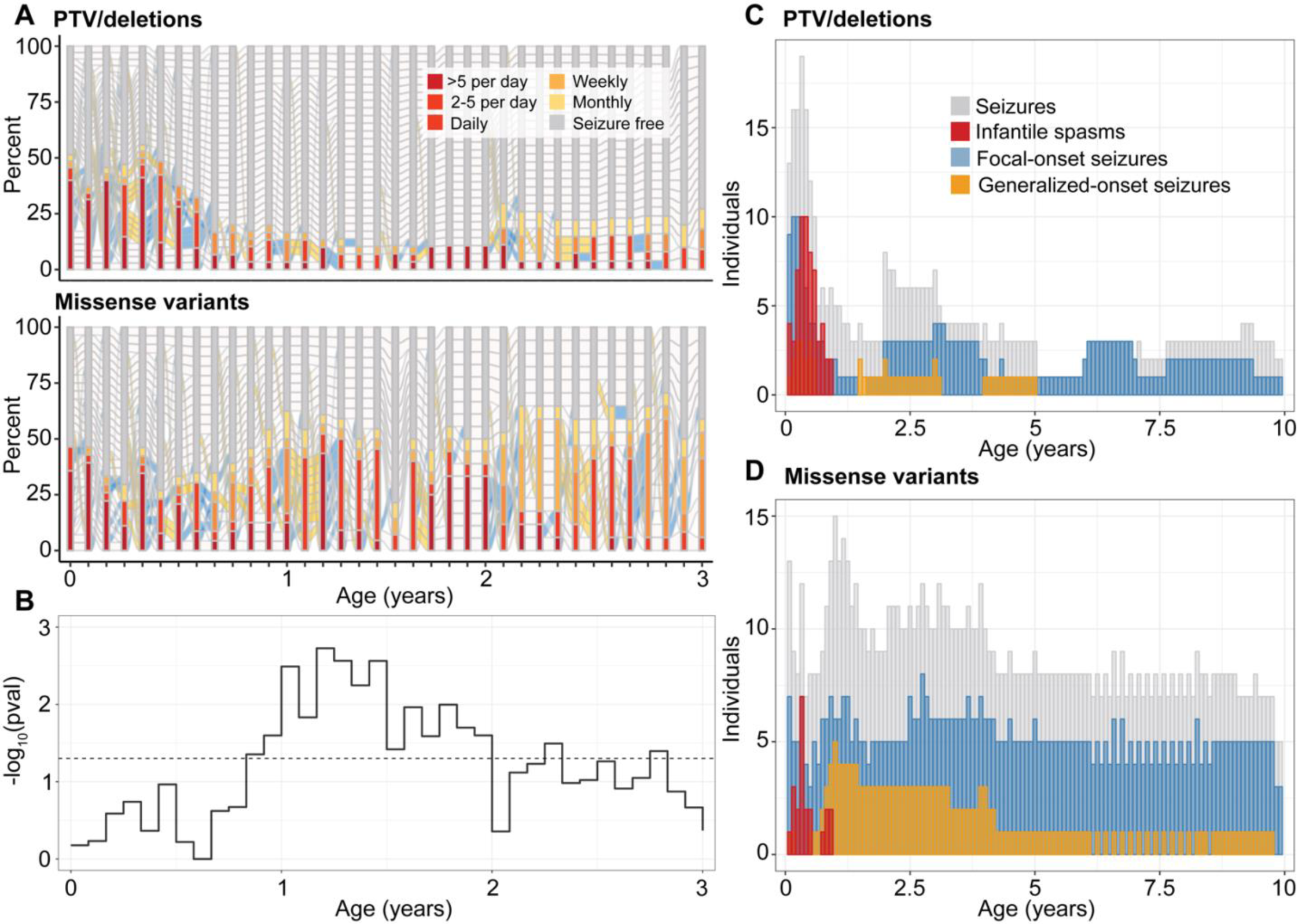
Difference in seizure histories between individuals with protein-truncating variants (PTV) and deletions versus missense variants in *STXBP1*. (A) Progression of epilepsy on a monthly basis in a subgroup of individuals with seizures at any point in the first 3 years of life (n=30 missense, n=39 PTV). **(B)** Longitudinal significance of the difference in the distribution of overall seizure frequencies in each respective month across the two subgroups (above). **(C)** Distribution of common seizure types across individuals with PTV/deletions, contrasted with **(D)** the distribution of seizure types across individuals with missense variants in *STXBP1* in the first 10 years of life.

Lastly, we compared epilepsy severity across the age span in individuals with recurrent missense variants with suspected dominant effects, including p.Arg406Cys/His/Ser (n=9), p.Arg292Cys/His/Pro (n=8), and p.Arg551Cys/His (n=3, **Supplementary Figure 1**). Individuals with p.Arg292Cys/His/Pro had slightly higher risk for neonatal seizures (n=4, p=0.04) and focal-onset seizures throughout the first two years of life. We also found a significant difference with frequent seizures between 12 and 18 months of life in two individuals with p.Arg551Cys/His, overlapping the period during which seizure remission was observed in >50% of the remainder of the cohort. When grouping the three recurrent variant hotspots (p.Arg406Cys/His/Ser, p.Arg292Cys/His/Pro, and p.Arg551Cys/His, n=20), the association was less robust with only an increase in tonic seizures between 7 and 8 months, which was primarily driven by two individuals with p.Arg292Cys/His/Pro variants. This may suggest that unique epilepsy signatures are associated with specific recurrent variants rather than with the broader variant group with suspected dominant-negative effects.

### Development in *STXBP1* is highly variable in the first five years of life, with emerging stratification by markers of disease severity

When analyzing milestones assessed in at least 20 individuals (**Figure 3A/B**), we found a range of percentages of achieved milestones, spanning from ability to use non-verbal communication (n=20) (f=1.00), to ability to roll over (n=121/135, f=0.90), ability to sit unsupported (n=96/133, f=0.72), ability to walk (n=86/151, f=0.57) and walk unassisted (n=45/99, f=0.45), to ability to use sentences and verbal communication (n=14/102, f=0.14). We analyzed the cumulative onset of commonly assessed milestones (**Figure 3A**), namely, the ability to control head posture, roll over, sit unassisted, grasp, walk unassisted, and initiate words and use verbal communication. Development across the domains were primarily within the first five years of life, and of individuals noted to achieve each respective milestone, the median age of head control was at 6.3 months (n=31, IQR 3 months – 1 year, 90% cumulative onset at 3 years), roll over at 7.6 months (n=103, IQR 5 months – 11 months, 90% cumulative onset at 1.2 years), sit unassisted (n=84, IQR 10 months – 1.4 years, 90% cumulative onset at 2.0 years), grasp (n=60, IQR 7 months – 1.0 years, 90% cumulative onset at 1.96 years), walk (n=81, IQR 1.5 years – 3.47 years, 90% cumulative onset at 5.3 years), and initiate words (n=22, IQR 1.6 years – 3.3 years, 90% cumulative onset at 1.2 years, **Table 2**).

**Figure 3.**
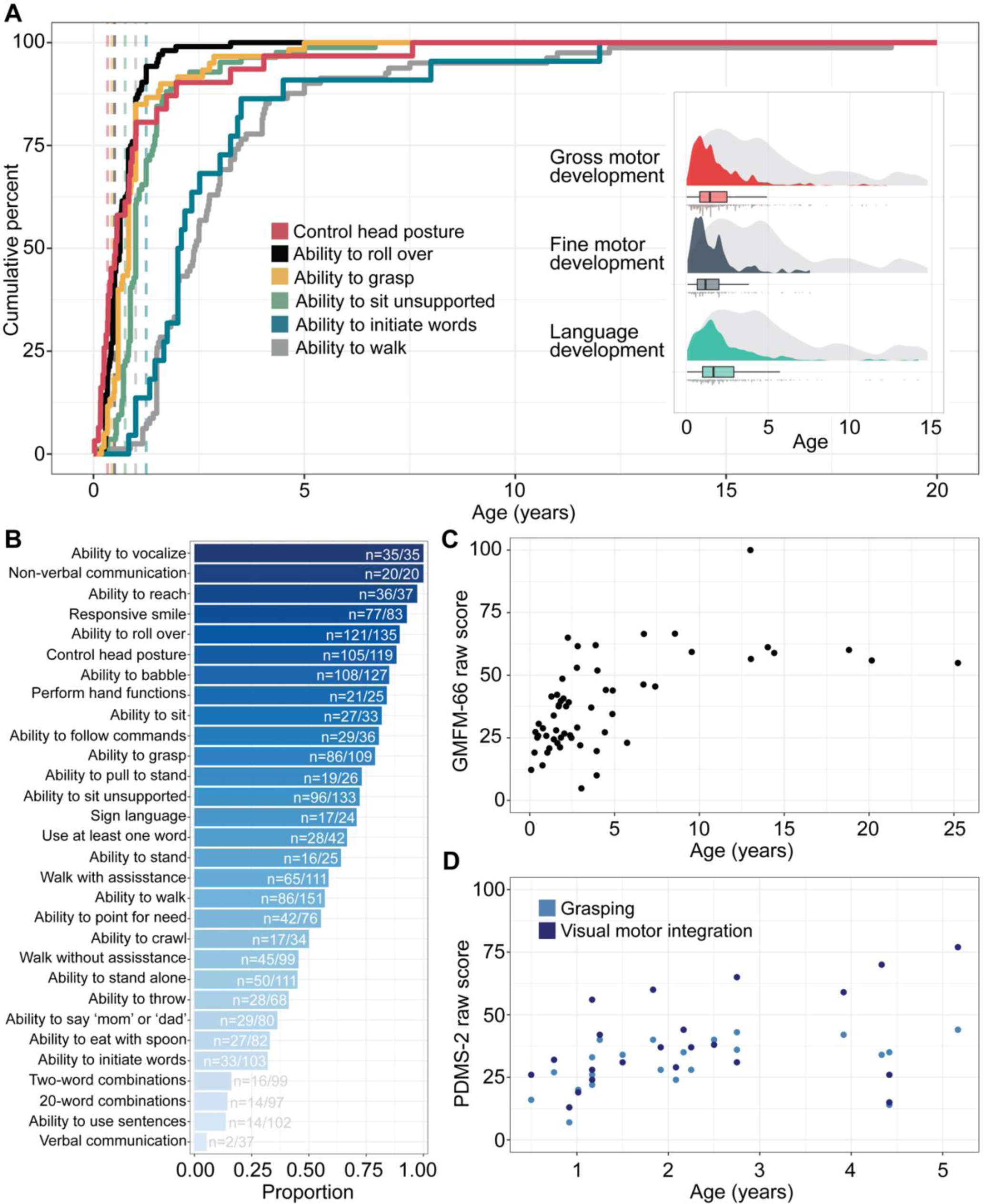
**Developmental outcomes and endpoints in *STXBP1*-related disorders. (A**) Cumulative milestone acquisition by age in 158 individuals with *STXBP1*. Dashed lines indicate the age at when 75% of typically developing children achieve each respective milestone as reported by the Center for Disease Control and Prevention (CDC). The distribution of gross motor, fine motor, and language development is primarily in the first 5 years of life (inset, showing age achieved indicated by each respective color versus age at which individuals were last assessed in grey). **(B)** Proportion of milestones achieved in the overall cohort, showing a wide range of percentages with verbal communication and language at the lower end. **(C)** Distribution of 57 GMFM-66-IS scores over time across 45 individuals. **(D)** Distribution of PDMS-2 raw scores for grasping and visual motor integration over time across 21 individuals.

Next, we assessed developmental trajectories using standardized outcome measures and classification scales (**Figure 3C/D; Table 2**). For gross motor classification, five individuals were Level I on the GMFCS-ER, 16 individuals Level II, 4 individuals Level III, 12 individuals Level IV, and 5 individuals had a Level V classification. Only 9 individuals were assessed for MACS to characterize ability to navigate and perform in daily life, for which no individuals had a Level I classification, four were Level II, two Level III, two Level IV, and one individual was Level V. Classification of language skills using the CFCS revealed that no individuals had a Level I classification, one was Level II, three level III, five Level IV, and 14 were Level V. Stratification of GMFM-66-IS scores by age ranges revealed a wide range of outcomes, with the greatest variability between the second and fifth year of life (median score of 35.8, IQR 25.75 – 46.05) and an observed ceiling of gross motor function outcome on the GMFM-66-IS in late childhood (median score of 58.9, IQR 54.9 – 61.2). When quantifying fine motor development using the PDMS-2 measure, we found that individuals with *STXBP1* had a range of scores for grasping and visual motor integration that mapped to an age equivalent of less than 14 months, even after 2 years of life (**Supplementary Figure 2**). The distribution of raw scores for PDMS-2 demonstrated a floor effect after the first year of life, when PDMS-2 percentiles in the cohort for visual motor integration dropped to below the first percentile and the overall range for grasping remained less than the 20th percentile.

Lastly, we aimed to map the range of developmental outcomes associated with markers of seizure severity, including subgroups stratified by the presence or absence of epilepsy or spasms and seizure offset versus ongoing seizures. We found that individuals with a current or past history of epilepsy and individuals with spasms had lower GMFM-66-IS scores than compared to individuals without any history of epilepsy (p=0.01) and without spasms (p=0.04), respectively (**Supplementary Figure 3**). We did not find a significant difference in GMFM-66-IS scores between individuals with ongoing seizures and individuals who were seizure free.

### Longitudinal forecasting of seizures reveals subgroups with different trajectories

We then characterized the range of epilepsy trajectories and identified subgroups defined by seizure predictability. The first aim was to delineate subgroups based on the variability of seizures. To this end, we developed a longitudinal seizure frequency forecasting model that quantified the likelihood of seizure frequency outcomes after the first year of life, for individuals with complete histories in the first year of life (n=78). Predictions compared to a distribution of randomized frequencies through permutation testing demonstrated “better than chance forecasting” across 87% of the cohort. The remaining individuals with unpredictable trajectories included primarily individuals whose seizures remained refractory throughout childhood but also individuals who had periods of seizure freedom despite predictions to have ongoing seizures.

We then compared the difference between predicted and actual seizure trajectories (**Figure 4A**) after the first year and derived a measure of variability across the cohort. This allowed us to characterize two subgroups: the “high-fidelity” subgroup included individuals with predictable trajectories (n=53), including 26 individuals who remained seizure free up until time of assessment, while the “low-fidelity” subgroup was defined by individuals with unpredictable trajectories (n=25), including the individuals whose seizures could not be forecasted better than chance (**Figure 4B**). This suggests that up to two thirds of individuals with *STXBP1* follow a predictable trajectory that can be reasonably inferred from the first year of life.

**Figure 4.**
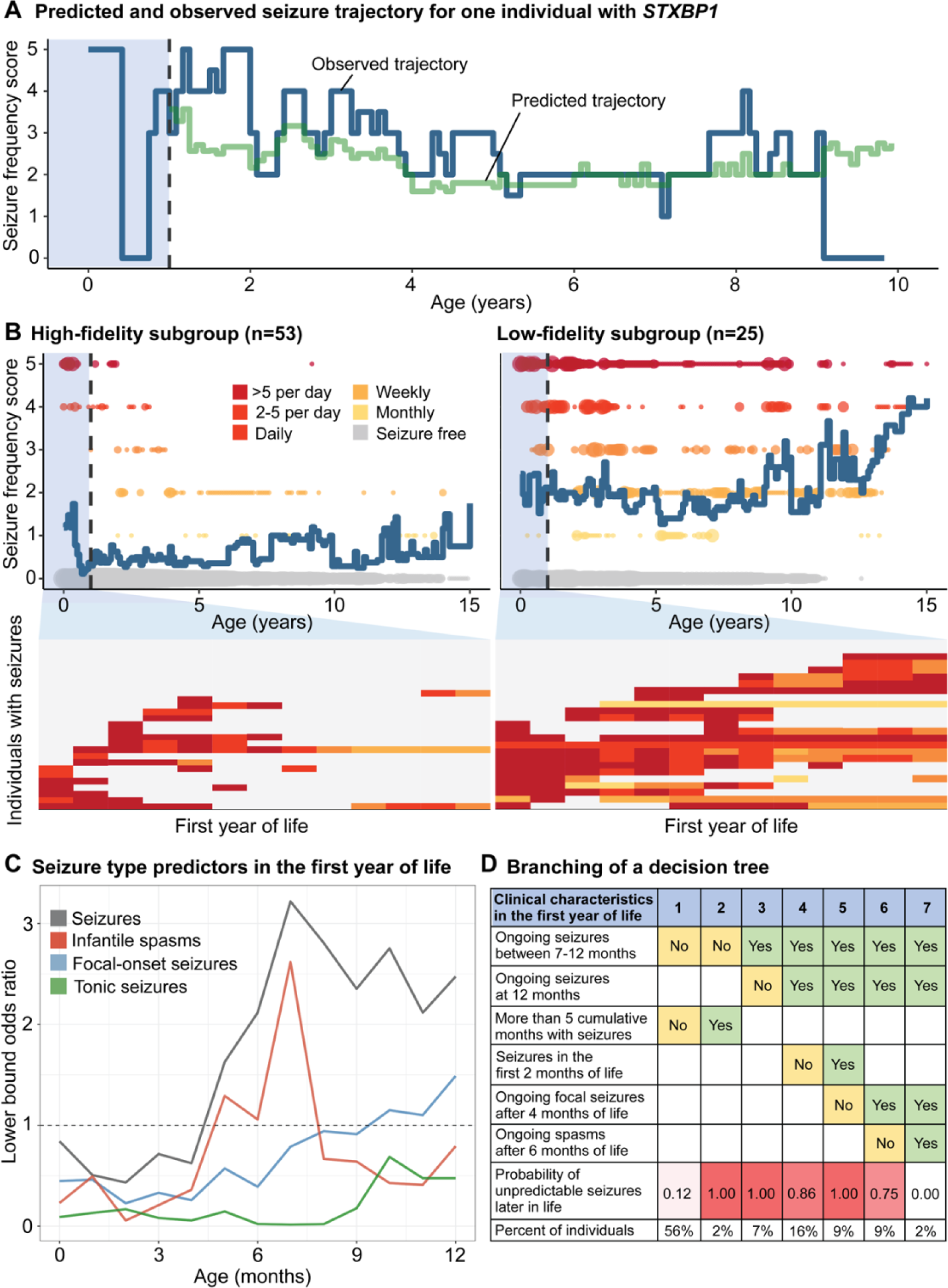
Longitudinal modeling of seizure histories in the first year of life in *STXBP1* revealed two subgroups with unpredictable and predictable epilepsy trajectories later in life. (A) Observed and predicted monthly seizure trajectory for one individual. The seizure history during the first 12 months were used to define a reference cohort of most phenotypically similar individuals. Seizure frequencies in the reference subgroup were then used to predict the individual’s trajectory. The difference in observed and predicted seizure frequencies across months was used to characterize the variability. **(B)** Variability was assessed across the cohort (n=78), with two subgroups: high-fidelity subgroup defined by predictable trajectories versus low-fidelity subgroup defined by unpredictable trajectories, showing seizure frequencies in the first year of life that resulted in high or low-fidelity predictions of longitudinal epilepsy trajectories later in life. **(C)** Seizure types leading to risk of having unpredictable seizures later in life. **(D)** Single decision tree, showing the branching of subgroups based on epilepsy characteristics or predictors for unpredictable seizure trajectories.

The second aim of this analysis was to identify epilepsy characteristics in the first year predictive of unpredictable seizures later in life. We found that features suggestive of a future predictable trajectory included neonatal or early infantile seizures that resolved by 7 months of life, or a shorter duration of seizures in infancy (**Figure 4C/D**). Particularly, individuals who achieved seizure freedom within a few months following onset showed a highly predictable seizure trajectory. This subgroup accounted for approximately 50% of individuals with epilepsy. We were also able to forecast future refractory seizures in individuals who had focal seizures, infantile spasms after 6 months, and ongoing seizures at 12 months of life, however this subgroup represented only 2% of the cohort.

Conversely, seizure characteristics indicative of an atypical, unpredictable course later in life included refractory epilepsy throughout the first year of life, especially between 7 to 12 months, and longer periods of consecutive months with seizures, indicating a more severe epilepsy subgroup within *STXBP1*. In contrast to the high-fidelity subgroup, 60% of the low-fidelity subgroup had ongoing epilepsy at 12 months of life. Furthermore, while there was no notable difference in the proportion of individuals with infantile spasms between subgroups (n=9/27 in high-fidelity, n=11/25 in low-fidelity subgroup), the longitudinal distribution of seizure frequencies after 6 months of life suggests that the differentiating factor was determined by ongoing and intractable spasms and seizures during this period. There was no significant difference in the variant spectrum between the high-fidelity (n=28 PTV/del, n=25 missense) and low-fidelity subgroups (n=13 PTV/del, n=9 missense, n=3 in-frame indel).

### Treatment effects in *STXBP1* vary over time and align with existing guidelines

We assessed existing treatment strategies and characterized the relative effectiveness of ASMs in our cohort. We reconstructed ASM histories across 307 total patient-years in a subgroup of 61 individuals with a total of 20 unique ASMs in addition to the ketogenic diet. The most frequently prescribed medications were levetiracetam (n=45, 99.5 patient-years), phenobarbital (n=37, 23.2 patient-years), topiramate (n=24, 31.3 patient-years), ACTH (n=21, 2.25 patient-years), and vigabatrin (n=20, 22.6 patient-years). The treatment landscape was dynamic across the age span, with phenobarbital, ACTH, and levetiracetam commonly used for treating neonatal and early infantile seizures, vigabatrin and levetiracetam in early childhood, and clobazam and the ketogenic diet preferred as treatment strategies later in life (**Figure 5A**).

**Figure 5.**
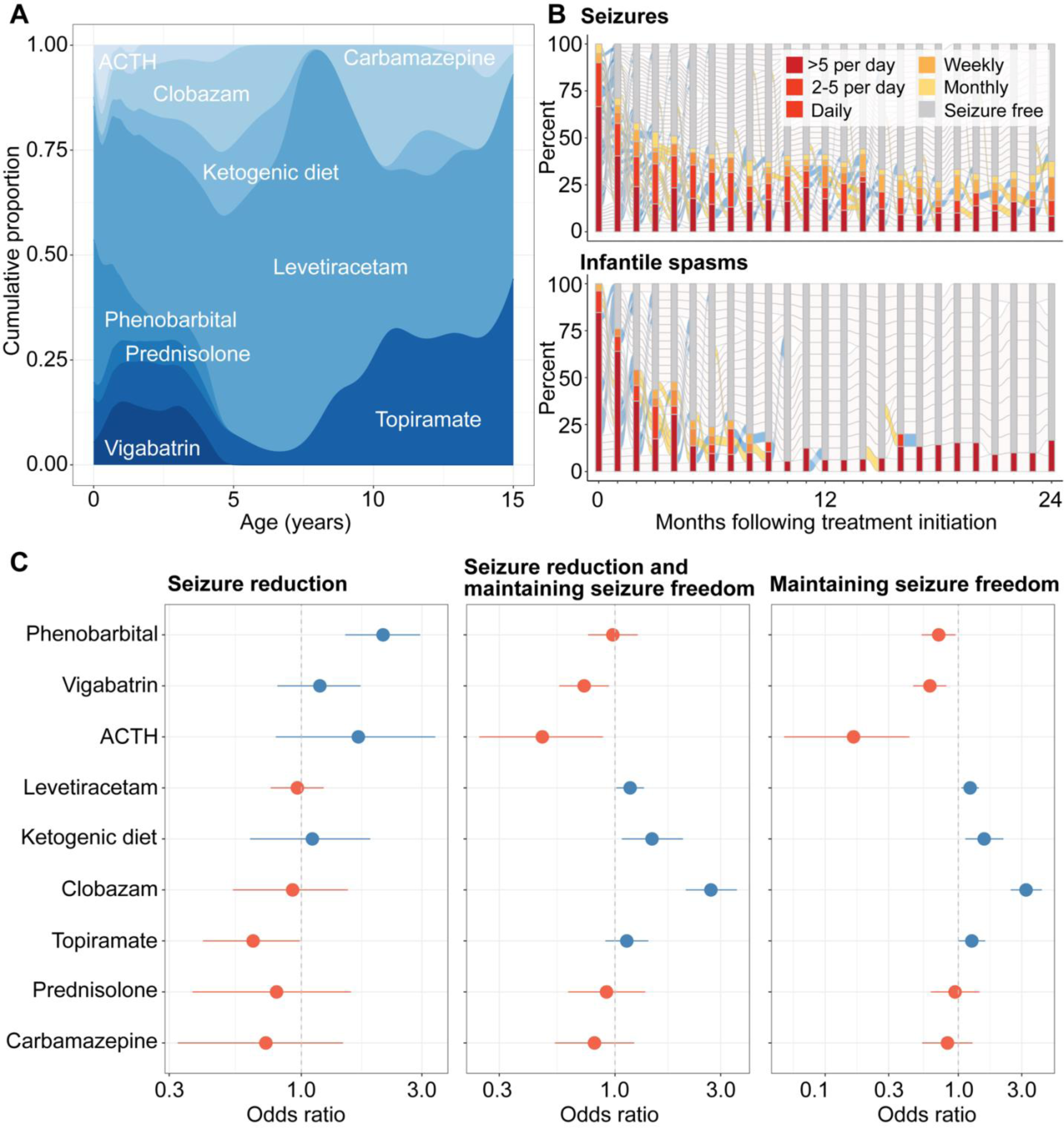
Characterization of treatment strategies and medication response in *STXBP1*-related disorders. (A) Anti-seizure medication landscape across 61 individuals with *STXBP1*, showing the relative density of ASMs used in at least 10 individuals across the age span. **(B)** Characterization of medication response over time following treatment initiation, showing the reduction in seizure frequencies on a monthly basis for seizure more broadly (top) and spasms (bottom). **(C)** Relative effectiveness of ASM for seizures vary across short-term and long-term treatment response, with phenobarbital as a superior ASM in reducing seizure frequency while clobazam and the ketogenic diet were effective in maintaining seizure freedom. Only medications used in at least 10 individuals are shown.

We found that the use of any ASM or combination of ASMs was able to reduce overall seizure burden by approximately 50% within 5 months following initiation. This effect was seen to a greater extent for treating spasms, in which 50% of individuals with spasms had offset within 3 months following ASM initiation (**Figure 5B**). When assessing seizure burden across epilepsy syndromes, we found a difference in medication response; individuals with Lennox-Gastaut Syndrome (LGS) and Developmental and Epileptic Encephalopathies (DEEs) (n=15) were significantly less likely to experience a reduction in seizure frequencies when compared to individuals with West Syndrome and individuals with other early onset epileptic encephalopathies (EOEE) (n=22) (**Supplementary Figure 4**). This points to differences in disease severity within *STXBP1*-related disorders.

We then analyzed the relative efficacy for both short-term and long-term response (**Figure 5C**). Phenobarbital was two-fold more likely to reduce overall seizures (n=37, OR 2.11, 95% CI 1.49-2.95), primarily driven by the offset of neonatal seizures in a subgroup of 21 individuals by 4 months of life. When accounting for periods of seizure freedom, we found that clobazam (n=14, OR 2.71, 95% CI 2.09-3.55) and the ketogenic diet (n=10, OR 1.47, 95% CI 1.07-2.03) were superior treatment strategies. The medication response in clobazam and ketogenic diet was seen to a greater degree when assessing for maintaining seizure freedom only, and topiramate emerged as another ASM with relative efficacy compared to other medication choices (n=22, OR 1.26, 95% CI 1.00-1.59). Accordingly, we derived a framework for quantifying longitudinal response to ASMs in *STXBP1*-related disorders from real-world data, providing a foundation upon which the efficacy of future therapeutic and treatment strategies can be evaluated.

### Virtual clinical trials demonstrate an age-dependent distribution of treatment success

Given the heterogeneity of seizure trajectories in *STXBP1*-related epilepsy, we identified optimal time windows during which a treatment effect would have the highest probability of being detected in a clinical trial. Virtual clinical trials using seizure frequency as the primary outcome resulted in wide range of trial success probabilities across the age span. We developed a measure for the probability of success in a virtual clinical trial: the Observed Frequency of Trial Success (OFTS), defined as the proportion of 1,000 simulated trials in which a significant difference (p<0.05) was detected. We were able to achieve the highest OFTS (range 0.87 to >.95) in early childhood between 8 months and 3.5 years of life (**Figure 6A**), with a wide overall range of probabilities from 0 to >0.95. While we found a second window between 7 and 10 years of life, this effect was driven by the repeated sampling of a limited number of individuals with ongoing refractory seizures during this period. When stratified by seizure type (**Figure 6B/C/D**), we found an optimal window starting at 3 months and a second window after 7 months for individuals with intractable spasms, 6 to 18 months and 2 to 3.5 years for focal-onset seizures, and two periods spanning from 8 months to just prior to 2 years and 2 to 3.5 years for tonic seizures. This demonstrates an age and seizure type-dependent landscape for optimal treatment windows.

**Figure 6.**
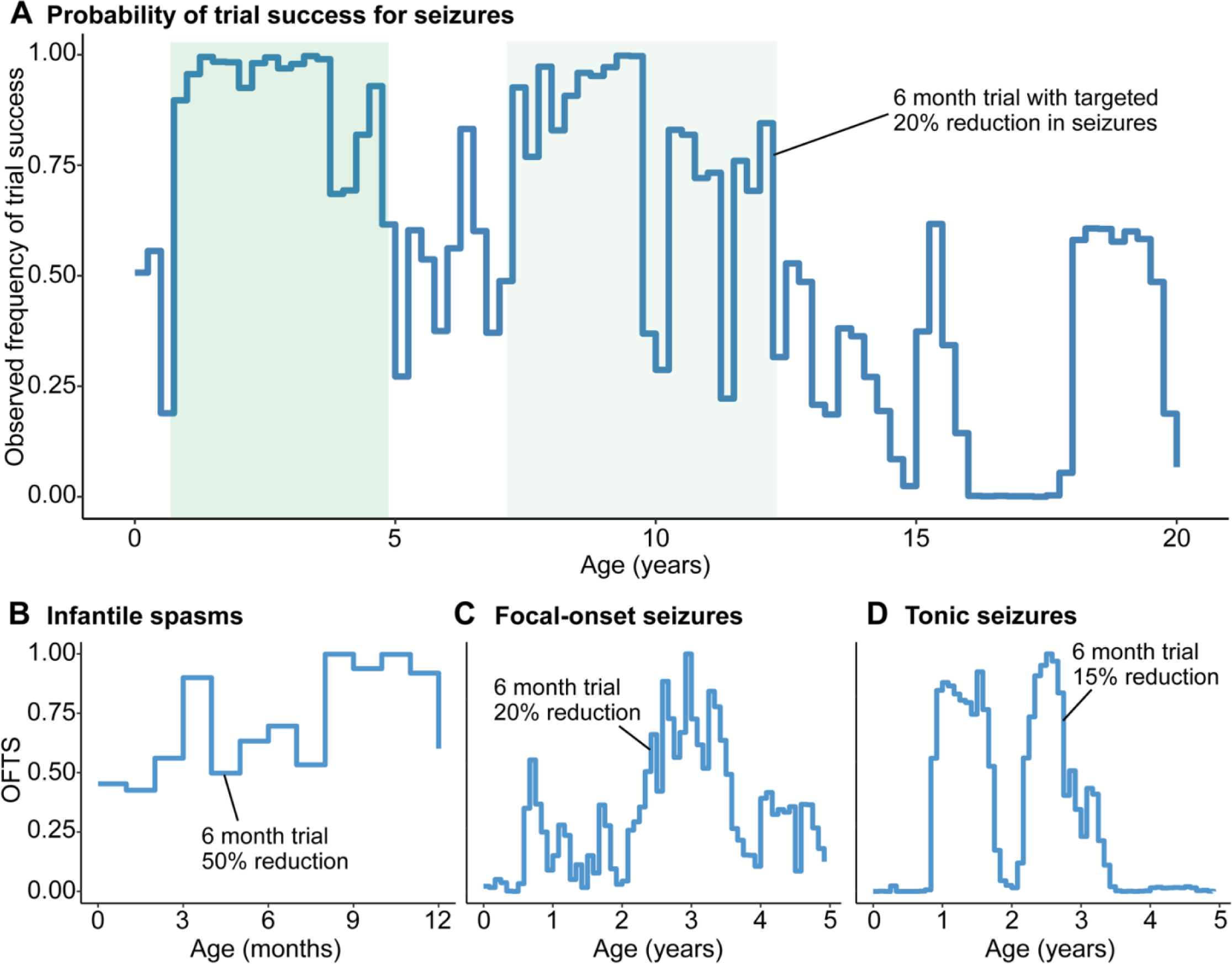
Virtual clinical trial framework for *STXBP1*-related disorders. (A) Virtual clinical trial using seizure frequency as the primary outcome, highlighting optimal age windows in green. The observed frequency of trial success (OFTS) is defined as the proportion of trials out of 1,000 virtual trials in which a significant effect can be detected if the trial were to be started at the respective month. A trial duration of 6-month versus 12-month period and the targeted seizure reduction ranging from 10% to 50% was selected based on the widest distribution of OFTS across the age span. **(B)** Virtual trials for targeting infantile spasms demonstrate an early window for optimal intervention starting at 3 months, and a second window is observed after 7 months for individuals with intractable spasms. **(C)** Virtual trials for targeting focal-onset seizures show an optimal window of 6 to 18 months and 2 to 3.5 years, although the OFTS was more variable. **(D)** Virtual trials for targeting tonic seizure, showing broad windows between 8 months to just prior to 2 years and 2 to 3.5 years.

## Discussion

Disease-causing variants in *STXBP1* lead to a heterogenous group of clinical presentations with varying disease severity and a wide range of outcomes. Understanding the natural history of *STXBP1*-related disorders, including long-term epilepsy trajectories and developmental endpoints, is critical to the development of clinical trials and delivery of novel therapeutic strategies. In our study of 162 individuals with *STXBP1*-related disorders, we found a heterogeneous pattern of seizures, especially in the first year of life and into early childhood, alongside high baseline variability in developmental domains including gross motor, fine motor, and communication abilities. Notably, we characterized epilepsy and development using validated outcome scales across the age span, demonstrating a dynamic disease trajectory that sets the groundwork for future prospective natural history studies and clinical trials.

### Seizures in *STXBP1*

Seizure onset is within the second month of life in 50% of individuals and in the first year of life in more than 80%. In contrast to other common genetic epilepsies, such as Dravet Syndrome,^38, 39^ the overall pattern of seizures in *STXBP1* is highly variable. While the first three months of life are characterized by the highest seizure burden, we found a heterogeneous interplay of seizure onset and offset in the first year of life, with few months in which seizures were present in more than 50% of the cohort, and a heterogeneous progression of different seizure types into early childhood. Focal-onset seizures were the most frequent type, followed by infantile and epileptic spasms. Neonatal and early infantile seizures in *STXBP1* may lead to earlier molecular diagnoses, which has important implications in prognosis and targeted care including early intervention for epilepsy management.

As previously recognized,^3^ we found that epilepsy histories diverge between individuals with missense variants compared to protein-truncating variants (PTV) and deletions. Individuals with PTV/del had increased risk of infantile spasms, while individuals with missense variants were more likely to have ongoing seizures later in life. When assessing recurrent missense variants with suspected dominant negative effects,^12, 14^ we found that individuals with p.Arg292Cys/His/Pro were at greater risk for neonatal seizures, and individuals with p.Arg551Cys/His were more likely to have ongoing seizures after 12 months of life, however these findings were driven by a limited number of individuals. Nevertheless, the identification and recognition of natural histories specific to underlying etiologies will be necessary in tailored prognostic care and in moving towards genotype-guided trials in the future.

### Developmental outcomes

With increased clinical genetic testing, an increasing number of individuals are recognized to have developmental delay and intellectual disability without epilepsy within *STXBP1*-related disorders.^1, 5, 8^ Our cohort included 30% individuals without seizures, pointing to the need to delineate developmental endpoints as an additional outcome measure complementary to epilepsy endpoints. However, unlike for well-studied neurological conditions such as cerebral palsy (CP), disease-specific outcome measures for *STXBP1* and most rare neurogenetic conditions currently do not exist. While outcome measures and classification scales developed for children with CP may be used in *STXBP1*-related disorders,^40^ future longitudinal research is needed to determine if their trajectory of development is like children with the broader diagnosis of CP, especially if motor outcome is to be used to determine the effectiveness of future clinical trials.

We found that development is variable in the first five years of life, with observed improvement in gross and fine motor development and acquisition of milestones during this window. This underscores the importance of intervention in early childhood to maximize development and achieve better long-term outcomes. The need to provide interventions in early childhood to affect motor development is well established in children with CP. During childhood development, gross motor skills typically plateau by 4–7 years of age,^41^ while GMFCS levels remain stable throughout childhood.^42^ However, the early years of life are an exception with less stability in GMFCS classifications before the age of 2 years.^19^ Our study shows that most individuals with *STXBP1*-related disorders attain gross motor milestones including ability to walk unassisted in more than 45% of individuals and reach GMFM-66-IS scores ranging from 50-75 after 5 years of life. The wide range of outcomes suggest that gross motor development can be considered a primary endpoint as a comprehensive and modifiable measure.

Fine motor development in *STXBP1* was more variable, as demonstrated by most PDMS-2 percentiles mapping to an age equivalent of less than 14 months. The assessment of fine motor function in children with *STXBP1* may be difficult due to the need to follow directions, which may be limited by language and cognitive deficits.^43, 44^ As families of children with *STXBP1* have reported difficulties with activities of daily living as a major concern,^18^ additional research to validate reliable fine motor function measures is necessary as potential treatments for *STXBP1* are developed in the future.

In addition, challenges in language and communication may be more prominent in *STXBP1*-related disorders compared to other genetic epilepsies. Families of children with *STXBP1* have identified expressive and receptive communication challenges as major symptoms which affect their lives.^18^ Our study demonstrates that the milestones with lowest frequencies achieved across the cohort are primarily regarding differences in language abilities, including the ability to use words or sentences in less than 25% of individuals. However, we find that almost all individuals assessed use non-verbal means of communication, highlighting that expressive communication is not limited to verbal communication. Additional longitudinal assessments of language function using more comprehensive scales such as the Observer-Reported Communication Ability (ORCA) Measure will be critical to examine the development of language skills in children with *STXBP1*.^45, 46^

Furthermore, current challenges in trials are the switching of developmental scales across the age span and floor effects, when existing scales cannot capture and differentiate the range of outcomes at the lower extremes.^47^ The use of raw scores rather than standard scores and testing children out the intended age range for outcome measures may be necessary to accurately measure children with *STXBP1*.^43^ The development of disease specific outcomes measures has been completed with some neurogenetic conditions such as *CDKL5* Deficiency Disorder and Aicardi Goutières Syndrome to avoid floor effects which could mask potential changes as the result of an intervention.^48, 49^

### Epilepsy variability and virtual clinical trials

Seizures in *STXBP1* are heterogeneous, and subgroups can be stratified based on the variability or “unpredictability” of seizures across the age span. Through a longitudinal seizure frequency forecasting model based on interrogation of epilepsy histories in the first year of life, we show that seizures can be forecasted better than chance in 87% of individuals. We quantified the degree of variability and identified two subgroups, namely individuals who had predictable versus unpredictable seizures later in life. The subgroup with high predictability included individuals with early infantile seizures that resolved within several months, which accounted for up to 50% of individuals with epilepsy. Conversely, individuals whose seizures diverged from forecasted trajectories constituted up to one third of individuals with *STXBP1*-related disorders, representing a high proportion of individuals with variable outcomes. Markers of epilepsy histories in the first year of life indicative of highly unpredictable seizure histories later in life included refractory epilepsy throughout the first year with ongoing seizures at 12 months. Understanding the clinical factors associated with specific trajectories is critical for clinical decision making, enabling personalized risk stratification and monitoring disease progression over time.^24, 33, 50–52^ Furthermore, longitudinal forecasting facilitates identification of atypical presentations which allow for variant prioritization in functional studies in addition to providing a prognostic baseline score for inclusion and exclusion of robust subgroups for future trials.

Seizure frequency is an objective measure that has thus far been used as the primary outcome when interpreting change from baseline in clinical trials in the epilepsies,^53, 54^ however the overall variability of seizures in a given patient cohort can decrease statistical power when evaluating treatment effect. Accordingly, we modeled virtual clinical trials using a synthetic control approach, leveraging real world data captured from routine clinical care to analyze the probability of trial success across the age span.^51, 55^ We demonstrate that the most optimal age range for detecting a significant treatment effect is between 8 months and 3.5 years of life in *STXBP1*-related disorders. The probability of success when targeting focal-onset seizures was more variable, and trials for spasms and tonic seizures revealed varying windows. In summary, we provide insight into the design of efficient and cost-effective trials in the future.

### Treatment efficacy

Anti-seizure medications (ASM) are the primary treatment strategy for epilepsy management, however, whether certain ASMs are effective have largely relied on provider experience. Our study demonstrates that a systematic approach to capture ASM response without *a priori* hypotheses from real-world data can be used as a guiding framework for characterizing the efficacy of current treatment strategies in *STXBP1*. We show that phenobarbital is effective for short-term treatment, while clobazam and the ketogenic diet are effective for long-term management. Stratification of treatment response across the age span, seizure types, and between subgroups will provide granular insight into subtypes in *STXBP1* that will have the greatest prospect of benefit in a future trial.

### Limitations

Our study presents findings on the natural history of *STXBP1*-related disorders, leveraging existing data captured from clinical care through a retrospective study. As our cohort represents a primarily pediatric cohort with the median age at last encounter at 9 years, future studies will benefit from understanding longitudinal epilepsy and developmental outcomes in adulthood. To this end, the outcomes for early to late childhood in our study likely include overrepresentation of a more severely affected subgroup for whom data was available, and accordingly, follow-up studies of the younger population in which the age at genetic diagnosis was earlier will be critical. Additionally, while our study demonstrated that clinical heterogeneity is associated with markers of disease severity, such as a greater degree of developmental delay in individuals with epilepsy and more variable seizures trajectories in individuals with refractory epilepsy early in life, we were not able to assess the impact of genomic background and variation contributing to the wide range of outcomes. Lastly, families of individuals with *STXBP1* have identified clinical features including gastrointestinal and respiratory symptoms that have been underrepresented in the existing literature and may serve as additional endpoints alongside measures for tremor and ataxia in *STXBP1*.^18^ Future prospective natural history studies may benefit from integration of a comprehensive range of symptom domains in affected individuals and expand upon the windows for which meaningful improvements can be measured.

### Conclusion

The clinical heterogeneity of *STXBP1*-related disorders has historically posed challenges in delineating longitudinal trajectories and endpoints. Here, we established a baseline to understand the disease trajectory in *STXBP1*-related disorders, which is critical for tailored care. We identified treatment windows and outcome measures that will facilitate interpretation of efficacy for future therapeutic strategies and developed a more precise understanding of the current landscape of treatment strategies. This information provides a framework focused on clinical trial readiness for assessing longitudinal phenotypes in *STXBP1*-related disorders.

## Data Availability

All data produced in the present study are available upon reasonable request to the authors.

## Acknowledgements

We would like to thank the individuals and families with *STXBP1*-related disorders who participated in our study.

## Abbreviations

ACMG: = American College of Medical Genetics and Genomics
ASM: = Anti-seizure medication
DEE: = developmental and epileptic encephalopathy
EOEE: = early onset epileptic encephalopathy
PTV: = protein-truncating variant

