## Supplementary Material for "Delineating clinical and developmental outcomes in *STXBP1*-related disorders"

1. **Supplementary Methods.** Longitudinal seizure frequency forecasting

2. **Supplementary Methods.** Framework for virtual clinical trials

3. **Supplementary Figure 1.** Epilepsy trajectories stratified by genetic variants in *STXBP1*

4. **Supplementary Figure 2.** PDMS-2 measure of age equivalency and percentiles

5. **Supplementary Figure 3.** Developmental outcomes stratified by seizure severity

6. **Supplementary Figure 4.** ASM response across epilepsy syndromes

7. **Supplementary Figure 5.** ASM response across seizure types

### Methods: Longitudinal seizure frequency forecasting and quantification of unpredictability

We developed a framework for longitudinal seizure frequency forecasting to characterize epilepsy progression and predictability. Standardized seizure histories were captured using a previously published scale derived from the Pediatric Epilepsy Learning Health System (PELHS)-championed framework for seizure severity, and seizure frequencies were grouped in the following categories: multiple daily seizures (>5 per day, SF score = 5), several daily seizures (2–5 per day, SF score = 4), daily seizures (SF score = 3), weekly seizures (SF score = 2), monthly seizures (SF score = 1), and no seizures (SF score = 0). All individuals with complete seizure histories in the first year of life (n=78 individuals) were included in the seizure frequency forecasting analysis.

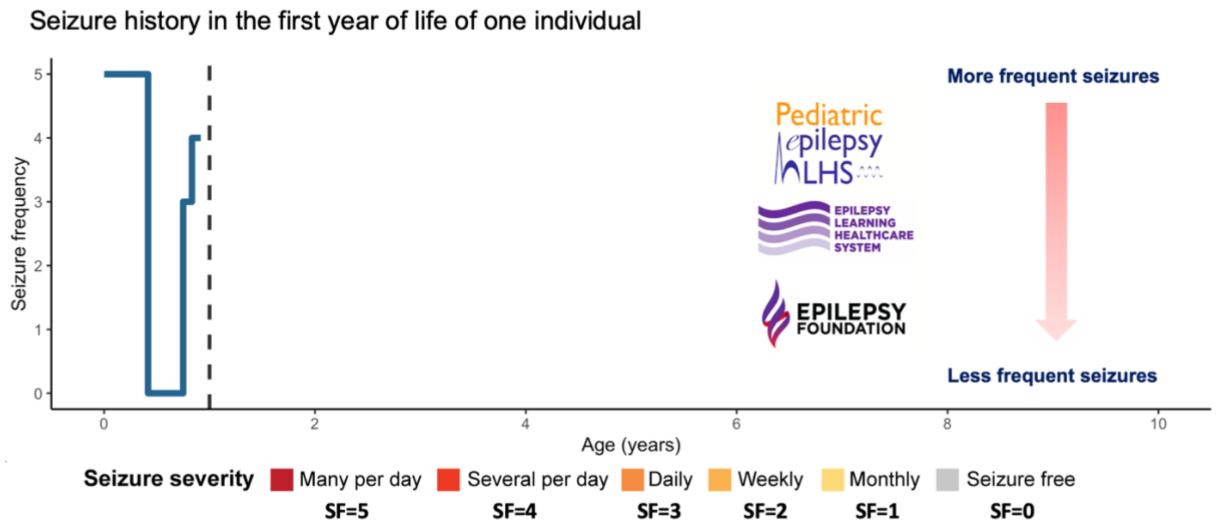


We compared seizure frequencies across monthly time intervals between all combinations of individual patient pairs during the first 12 months of life, when seizures are the most prominent in *STXBP1*. Based on the framework for semantic similarity analysis to measure phenotypic resemblance using clinical features (Galer et al., 2020; Helbig et al., 2019), we derived a complementary method to measure phenotypic similarity based on epilepsy histories, using monthly seizure frequency captured in the PELHS framework as comparisons (**see below**).

First, we calculated the information content (IC) of each seizure frequency (SF) category in each month (e.g., SF score = 3 during Month 8 of life). The IC was defined as the -log2 of the frequency of individuals in the SF category during the respective month. For example, if 20% of the cohort had at least daily seizures during the 8th month of life, then the IC for SF score = 3 was defined as -log2(0.20) = 2.32. Accordingly, the higher the frequency of individuals with a certain seizure frequency in a specific month, the less informative that seizure frequency for the respective month. In contrast, if only 5% of individuals had daily seizures at 7 years of life, an SF score = 3 would be more informative, and thus weighed more when deriving phenotypic resemblance.

To quantify phenotypic resemblance between two individuals, we summed the minimum IC overlap of each month across all overlapping months. As in the semantic similarity analysis, the rationale was to derive a higher similarity measure for individuals with more rare and thus more informative, or distinguishing seizure frequencies. For example, if two individuals had a less frequent seizures across many months, the derived similarity measure would be higher than two individuals with the same seizure frequency across fewer months and higher than two individuals with a seizure frequency that was more common in the overall cohort. To correct for varying observation times between patient pairs, we adjusted the cumulative IC score by dividing by the number of overlapping months of comparison for each respective patient pair.

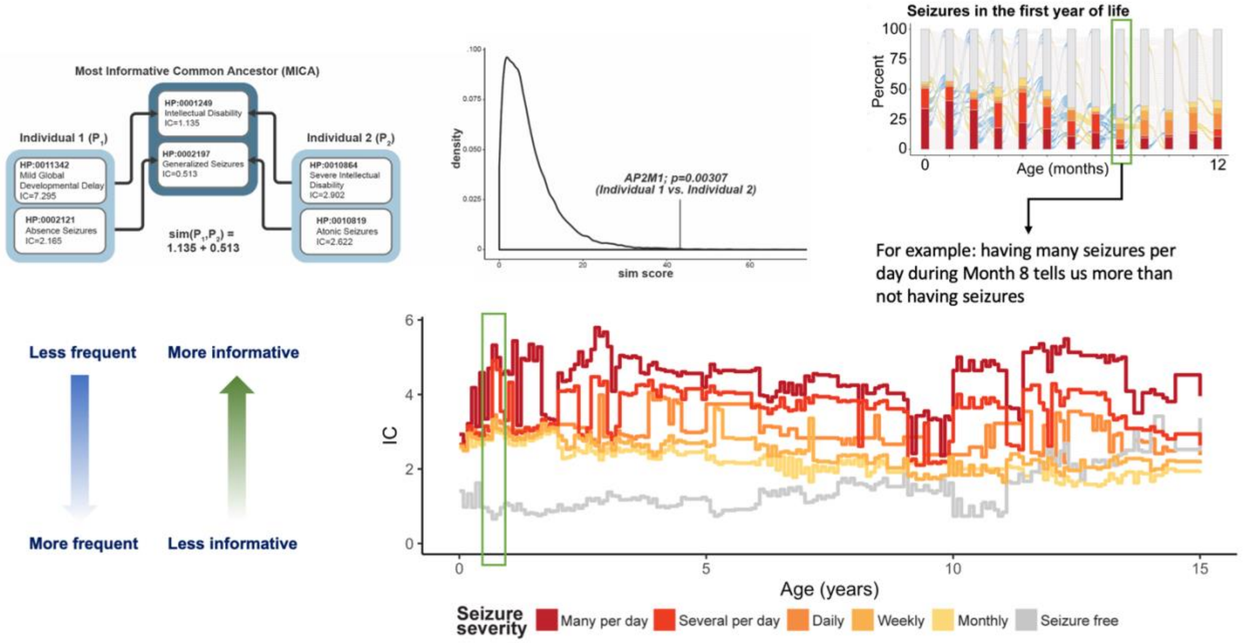


After deriving phenotypic similarity scores for all patient pairs in the first year of life, for each individual, we then identified 10 distinct individuals in the cohort that were the most similar phenotypically based on seizure frequencies across the age span, which was defined as that individual's reference cohort.

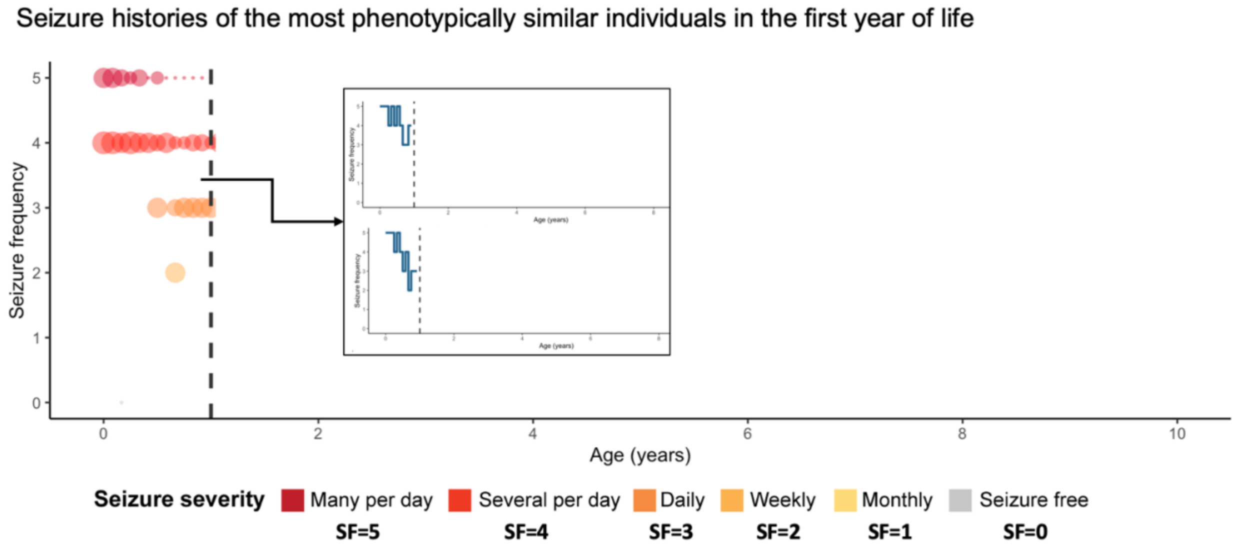


Then, with the known epilepsy histories in the reference cohort, including seizures after 12 months of life, we predicted each individuals' epilepsy trajectory after the first year, taking the median of the distribution of seizure frequencies for each month in the reference cohort, representing a distribution of likelihood of possible seizure frequency outcomes, as the predicted trajectory.

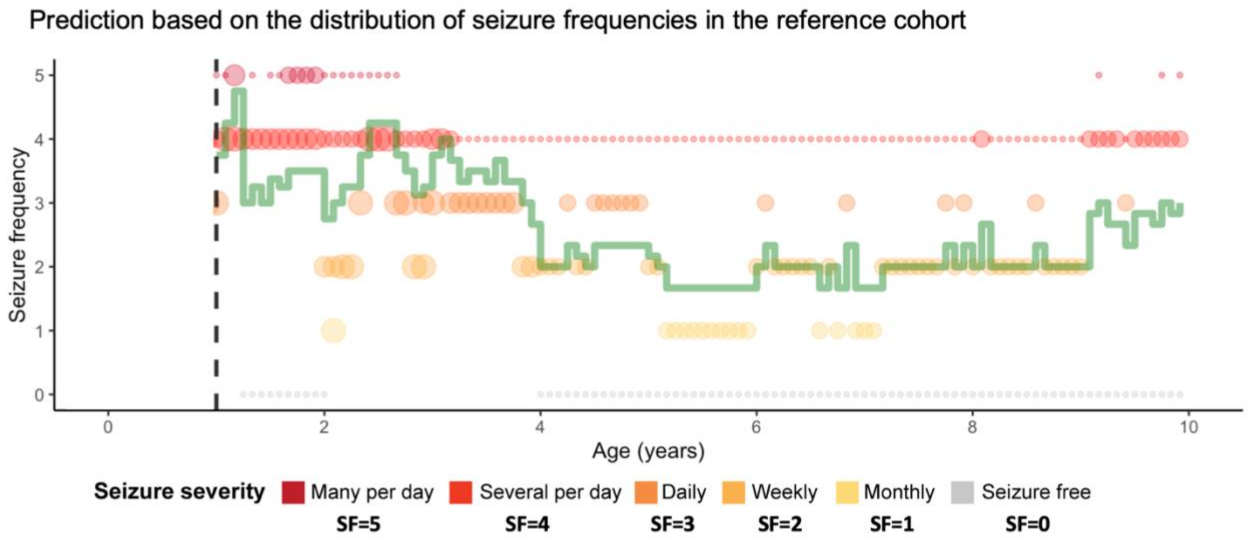


The predicted (example shown below in green) and actual seizure trajectories (example shown below in blue) after the first year of life was compared and assessing the cumulative difference enabled us to characterize subgroups defined by a high or a low difference between the forecasted and actual seizure frequencies, which we defined as a measure of epilepsy unpredictability. The grouping was performed using the k-means algorithm. For each individual, forecasted seizure frequencies were compared to a distribution of randomly generated seizure frequencies and permutation testing of 100,000 for each individual estimation allowed us to evaluate whether the predicted trajectory was better than chance.

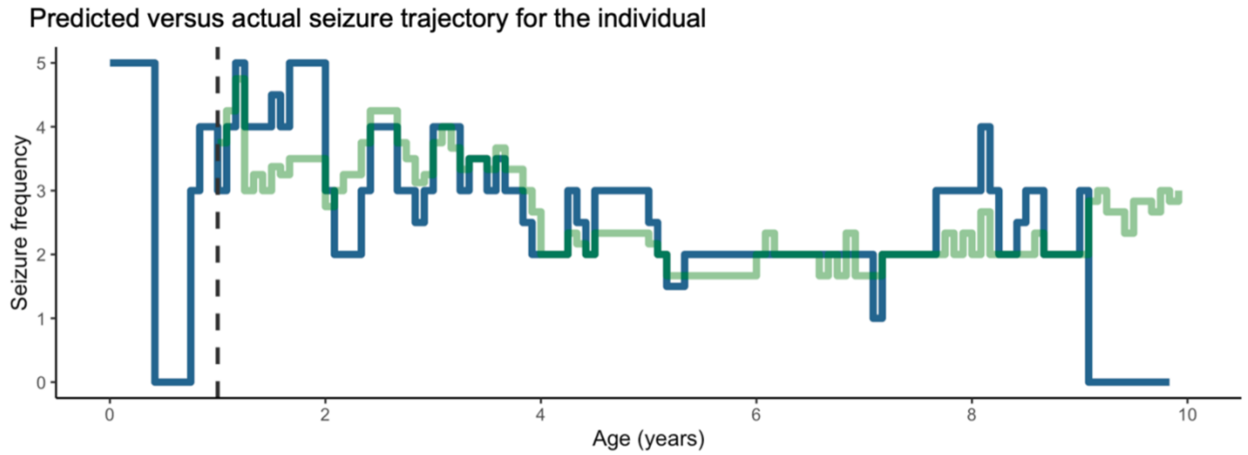

**Methods: Framework for virtual clinical trials**

Given the heterogeneity of seizures in *STXBP1*-related disorders, we aimed to identify time windows during which a treatment effect would have the highest probability of being detected in a clinical trial. We derived a framework for virtual clinical trials, randomly sampling 20 individuals with ongoing seizures and simulated a 6-month and 12-month period of 10%, 15%, and 20% seizure reduction.

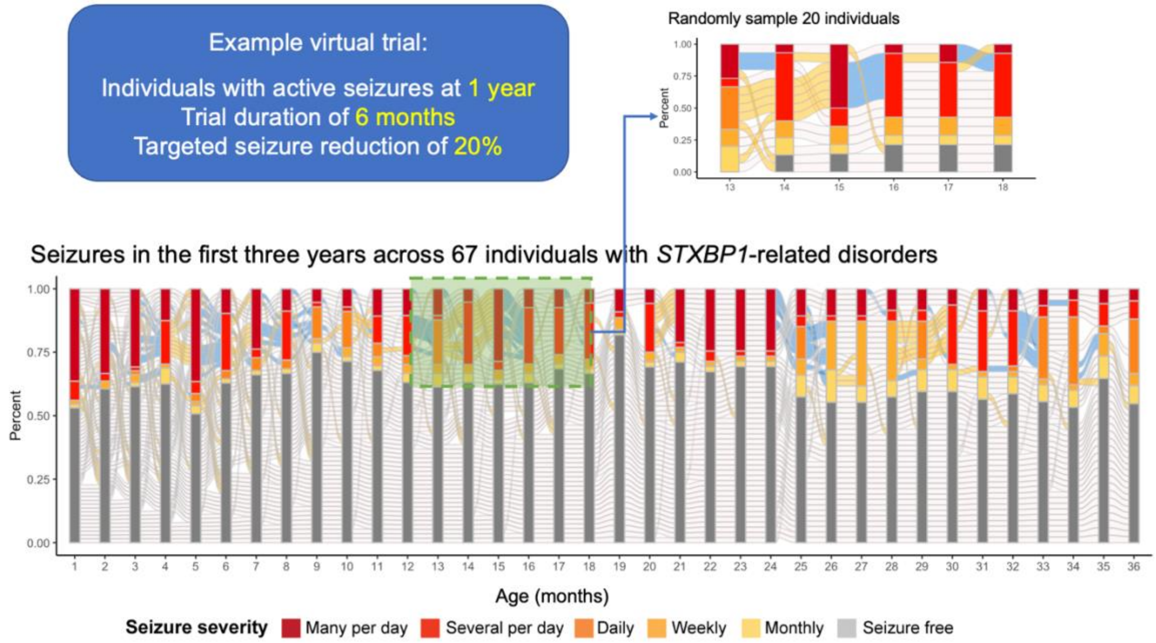


The percent reduction in seizures was calculated based on the cumulative sum of seizure frequencies at the start of the window, and the simulated treatment effect across each trial window was performed by decreasing seizure frequencies across the trial period for this cohort to create a synthetic treatment cohort.

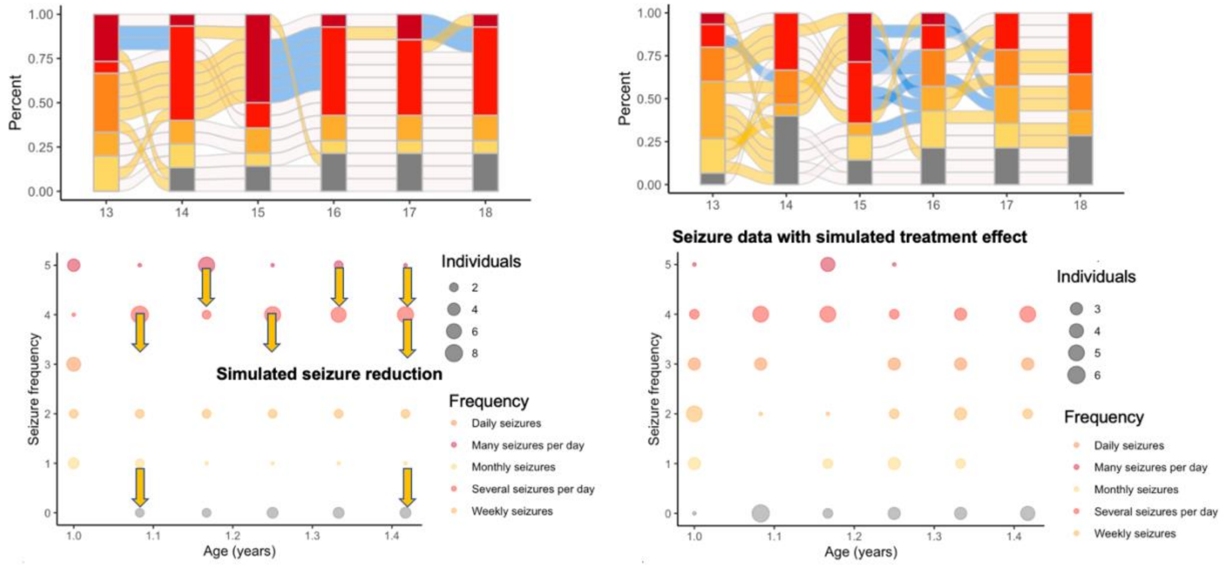

We used the synthetic control method to evaluate the significance of the simulated effect, comparing the distribution of seizure frequencies following simulated reduction of seizures against the observed, natural history distribution of frequencies in the sampled individuals using the Wilcoxon rank sum test.

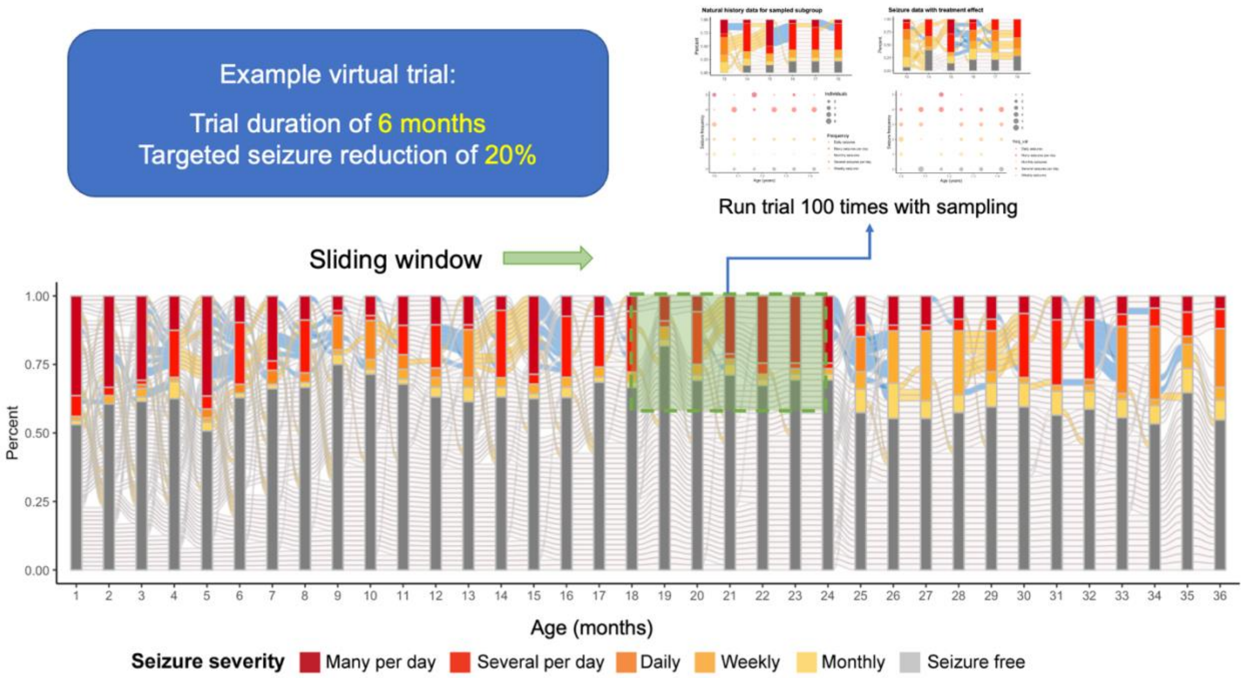


We ran 1,000 simulated trials for each month across the age span, shown above an example 6-month trial starting at 18 months of life. For each trial window, the Observed Frequency of Trial Success (OFTS) was defined as the proportion of trials out of 1,000 in which a significant effect could be detected. For analyses across seizure types, we chose to include the trial duration (6-month versus 12-month period) and targeted seizure reduction (10%, 15%, 20%, or 50%) based on the distribution of OFTS that resulted in the widest range, spanning from poor probability of trial success to high probability across the age span. This approach enabled us to identify optimal windows during which a treatment response would most likely be observed in a real-world trial when using seizure frequency as the primary outcome measure.

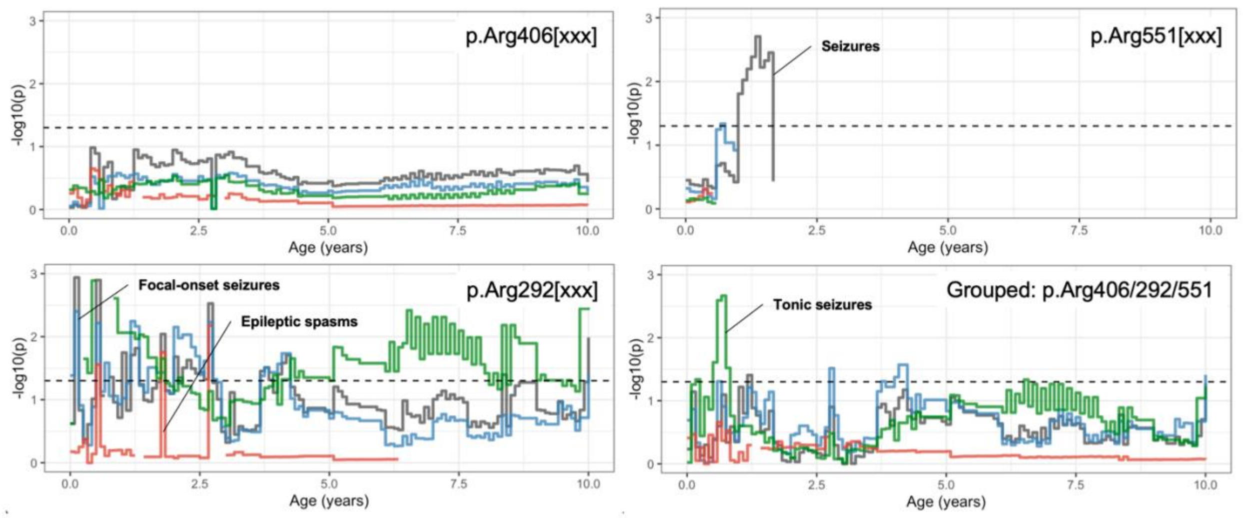


**Supplementary Figure 1. Epilepsy trajectories stratified by genetic variants in *STXBP1.*** We compared distributions in seizure frequencies across the age span in individuals with recurrent missense variants with suspected dominant effects, including p.Arg406Cys/His/Ser (n=9), p.Arg292Cys/His/Pro (n=8), and p.Arg551Cys/His (n=3).

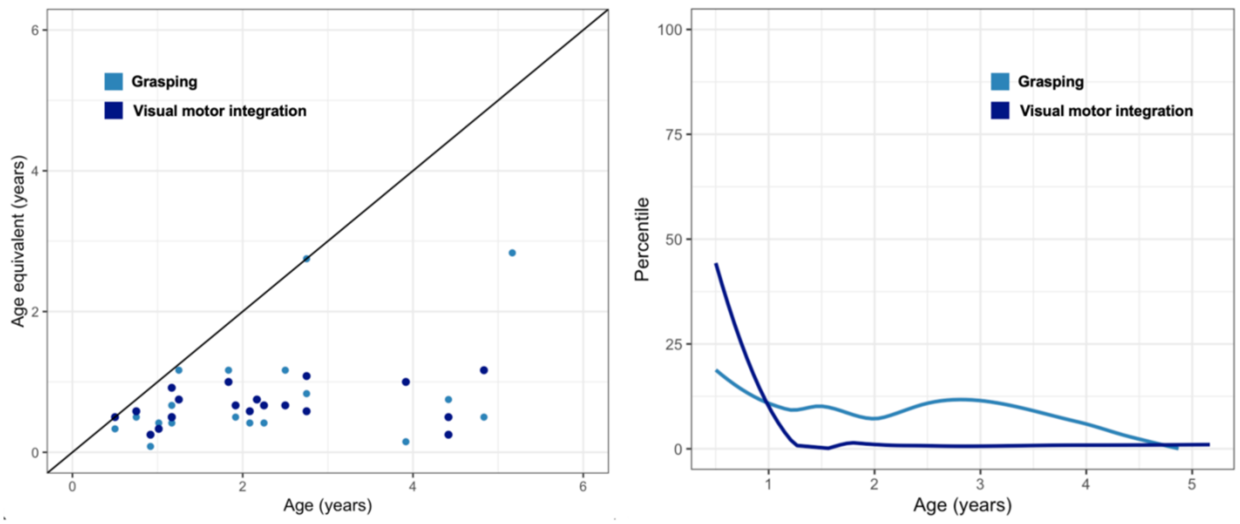


**Supplementary Figure 2. PDMS-2 measure of age equivalency and percentiles.** We assessed the range of PDMS-2 raw scores for grasping and visual motor integration, showing the age equivalent versus age at assessment (left) and corresponding percentiles across the age span (right).

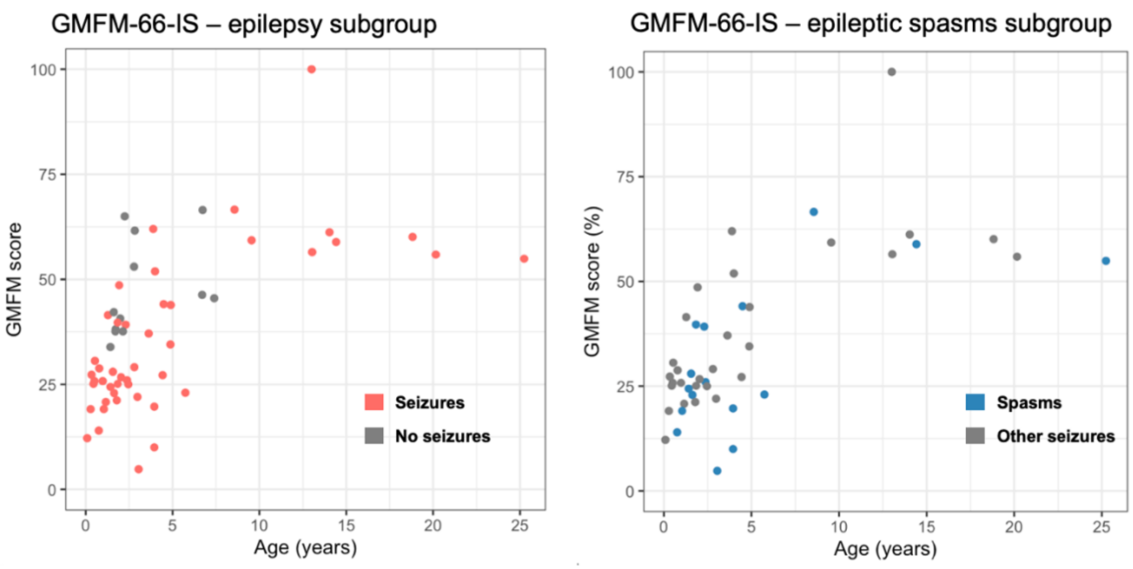


**Supplementary Figure 3. Gross motor developmental outcomes across epilepsy subgroups.** We assessed the range of GMFM-66-IS raw scores, stratified by individuals with seizures versus without seizures and individuals with spasms versus with other seizure types. The Wilcoxon rank sum test was used to determine statistical significance.

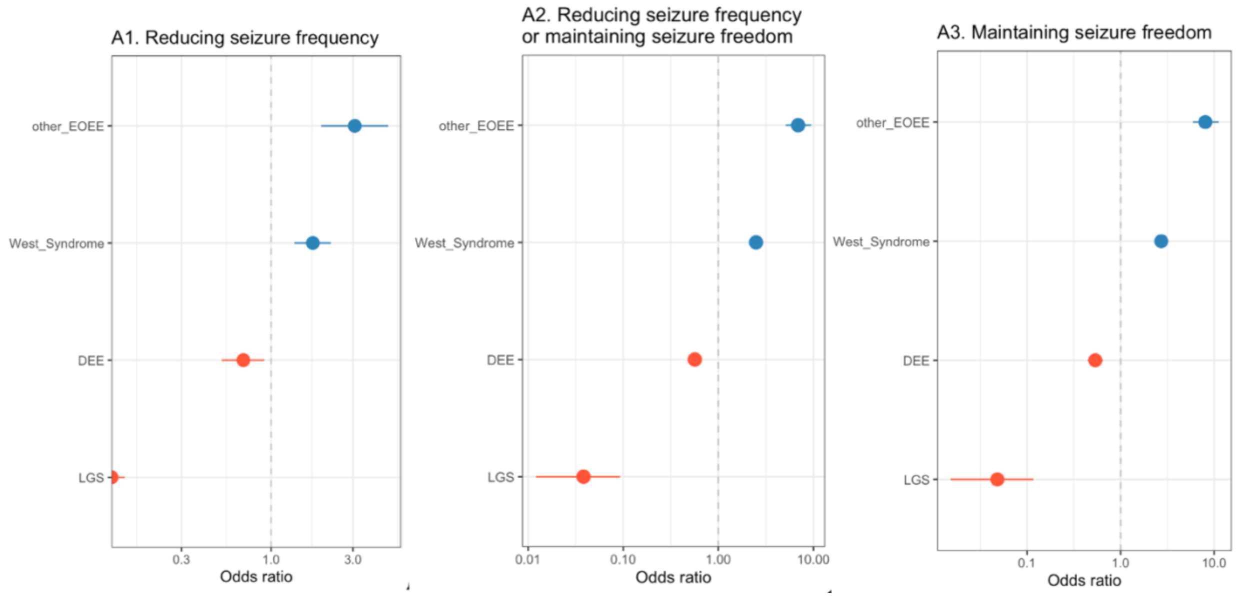


**Supplementary Figure 4. ASM response across epilepsy syndromes.** Medication response across epilepsy syndromes: West Syndrome, Developmental and Epileptic Encephalopathies (DEE), Lennox-Gastaut Syndrome (LGS), and Early Onset Epileptic Encephalopathies (EOEE). Odds ratios indicate relative the effectiveness of any anti-seizure medications (ASM) or treatment strategy in reducing seizure frequency and/or maintaining seizure freedom.

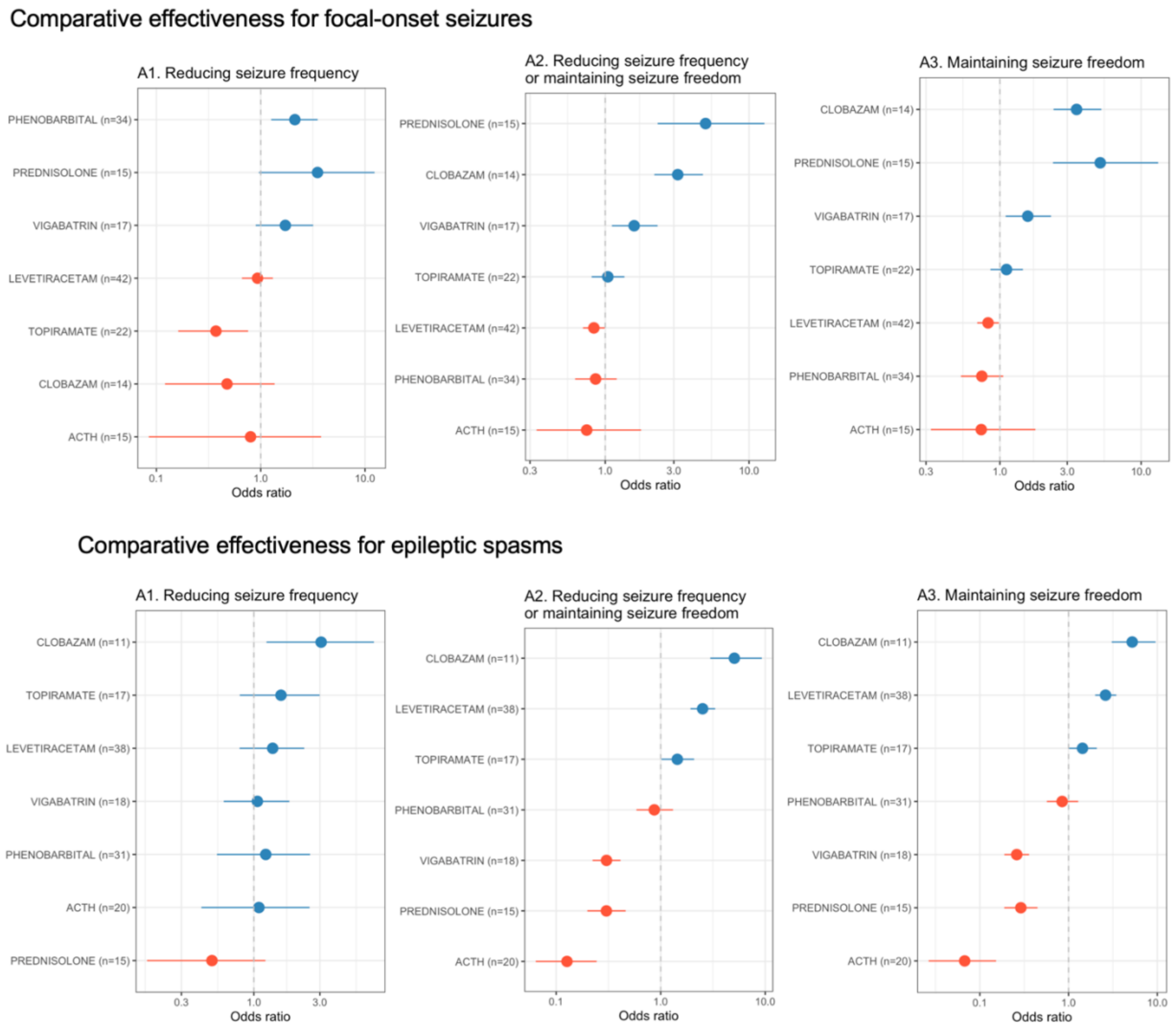


**Supplementary Figure 5. ASM response across seizure types.** Comparative effectiveness of anti-seizure medications stratified by individuals with focal-onset seizures and epileptic spasms, for short-term response in reducing seizure frequency (A1), in addition to long-term response in reducing seizure frequency or maintaining seizure freedom (A2) and maintaining seizure freedom (A3).
